# The cellular neighbourhoods of adult gliomas and their clinical associations

**DOI:** 10.64898/2026.11.24.26368523

**Authors:** Anna Mathioudaki, Samuel Rutz, Zaira Seferbekova, Gleb Rukhovich, Michael Ritter, Ekaterina Popova, Domenico Calafato, Sophia Schinkewitsch, Felix E. Hinz, Philipp D. Mahlknecht, Franziska M. Ippen, Heng Luo, Nela R. Koeberer, Nina Wilhelm, Michele Bortolomeazzi, Yiheng Tang, Areeba Patel, Pooja Sant, Jan-Philipp Mallm, Duncan T. Odom, Christel Herold-Mende, Miriam Ratliff, Nima Etminan, Sandro Krieg, Lukas Bunse, Michael Platten, Wolfgang Wick, Antje Wick, Andreas von Deimling, Sascha Dietrich, Felix Sahm, Abigail K. Suwala, Moritz Gerstung

## Abstract

Glial tumours are the most common primary malignant brain tumours in adults and comprise molecularly and histologically distinct entities with diverse outcomes. Despite this diversity, they arise within a shared anatomical environment, suggesting that common organisational principles may shape tumour heterogeneity. However, how cellular states are organised across gliomas and whether this organisation relates to clinical outcomes remains unclear. To that end, we generated a single-cell resolution spatial transcriptomic atlas of 2.6 million cells from 275 patients spanning IDH-wildtype glioblastoma, IDH-mutant and chromosome 1p/19q codeleted oligodendroglioma, IDH-mutant astrocytoma and ependymoma. Across diagnoses, we show how heterogeneous tumour cell states are spatially organised into recurrent neighbourhoods with distinct immune and glial composition, which in turn assemble into a higher-order architecture. Among these cellular neighbourhoods, some, such as the astrocyte-like (AC-like), are conserved across entities, whereas others segregate by clinicopathological parameters. AI-powered histopathology further reveals selective association of neighbourhoods within histological clusters, providing a link between molecular spatial organisation to routine H&E morphology. In astrocytoma, neighbourhood abundance is associated with patient survival independently of WHO grade, with AC-like enrichment linked to favourable outcomes. Same-cell high-plex spatial transcriptomics and multiplexed protein imaging in an independent astrocytoma cohort validated these neighbourhoods and further confirmed enrichment of oligodendrocytes within the TBI neighbourhood. Collectively, our findings reveal that glial tumours are spatially encoded through recurrent multicellular neighbourhoods across different molecular entities, showing clinical relevance beyond conventional histology and providing a foundation for analogous investigation across cancer types.

## Main

Despite decades of research, brain tumours remain among the deadliest cancers. Glial tumours, which are the most common primary malignant brain tumours, encompass a biologically and clinically diverse group of central nervous system (CNS) tumours. To account for this diversity, the current World Health Organisation (WHO) classification integrates histopathological features, DNA methylation profiling, and defining genetic alterations to stratify patients into distinct diagnostic groups ^1–3^. Even with this refined molecular taxonomy, clinical outcomes remain highly variable within diagnostic categories, motivating efforts to understand the cellular organisation of tumour tissue as a complementary axis of glioma biology.

Single-cell transcriptomic studies have identified recurrent malignant cell states – including astrocyte-like (AC-like), oligodendrocyte-progenitor-like (OPC-like), neural-progenitor-like (NPC-like), and mesenchymal-like (MES) states – within specific glioma subtypes, most notably glioblastoma ^4–11^. However, whether these states recur across the full spectrum of glioma entities remains unclear; distinguishing conserved from entity-specific features is therefore essential for understanding glioma heterogeneity. Similarly, resolving how tumour phenotypes are spatially organised within their shared anatomical environment could reveal organisational principles not captured by cell-state composition alone. Spatial omics offer the opportunity to resolve this organisation *in situ*, directly linking transcriptional identity to tissue architecture and the local microenvironment ^12–15^. Recent work using such methods has shown grade-dependent patterns of high-level tissue architecture in isocitrate dehydrogenase (IDH)-mutant gliomas, shifting from location-driven structure in low-grade tumours to hypoxia-driven organisation resembling glioblastoma in high-grade tumours ^16^. This suggests that tissue-level architecture may carry clinically relevant information. However, these studies lack single-cell spatial resolution, precluding identification of specific malignant states and cell–cell interactions underlying these spatial patterns, a gap that advancements in single-cell spatial profiling can now resolve ^17^.

Here, we apply a multimodal approach – combining single-cell spatial transcriptomics, DNA methylation and genetic profiling, multiplexed protein imaging, and computational histopathology – to systematically characterise recurrent tumour cell states and their spatial organisation across adult gliomas. To assess how broadly such organisational principles are shared, we study a diverse set of glial tumours. These include IDH-wildtype glioblastoma, IDH-mutant astrocytoma, chromosome 1p/19q-codeleted oligodendroglioma, thought to originate from neural stem or glial progenitor cells ^1^, alongside ependymomas, which are thought to arise from ependymal cells lining the ventricular system ^18–22^. By integrating these complementary modalities, we investigate how tumour-intrinsic programs and the local microenvironment jointly shape glioma tissue architecture. We further assess the clinical relevance of this spatial organisation for patient stratification.

### High-resolution single-cell spatial profiling across a diverse cohort of adult gliomas

To comprehensively characterise the cellular and transcriptional heterogeneity and recurrence patterns of tumour cells and the tumour microenvironment (TME) across adult gliomas, we assembled a cohort of 275 patients, from whom 282 tumour samples and 309 tissue cores were obtained for spatial transcriptomic profiling (**Figure 1A, B, C; Supplementary Table 1**). The cohort spans adult-type diffuse gliomas, including IDH-mutant (mut) and 1p/19q-codeleted oligodendroglioma (n=27), IDH-mutant astrocytoma (n=170), IDH-wildtype (wt) glioblastoma (n=24), as well as ependymal tumours, comprising subependymoma (n=15), myxopapillary ependymoma (n=19), and spinal ependymoma (n=20). One oligodendroglioma with sarcomatous features (oligosarcoma) showed no 1p/19q codeletion by copy-number analysis; oligosarcomas can retain 1p/19q loss of heterozygosity in a copy-number-neutral state ^23^. Of the patient samples profiled, 205 were primary and 74 were recurrent, including 5 patients with matched primary and recurrent samples. For a subset of patients, multiple tumour cores per tumour were included, allowing direct comparison of transcriptional variation across distinct regions within the same tumour (**Extended Data Figure 1**). As the majority of intra-patient comparisons showed consistently higher similarity than inter-patient comparisons, cores from the same patient were aggregated for downstream analyses.

DNA methylation profiling and panel sequencing data for recurrent driver gene mutations were available as part of routine diagnostic testing (**Figure 1C**). DNA methylation-based classification further refined these diagnoses, stratifying tumours into 13 molecular subtypes, including Receptor Tyrosine Kinase class I (RTK I), RTK II, and Mesenchymal (MES) glioblastoma subclasses (**Figure 1B**). Within astrocytomas, methylation-based classification distinguishes low-grade from high-grade molecular subtypes beyond histology; given the established clinical relevance of this distinction, we treat low-grade (n = 103) and high-grade (n = 69) astrocytoma as separate groups throughout this study ^2^. Two patients contributed samples to both groups, as their tumours progressed from low-grade to high-grade astrocytoma at recurrence.

Tissue cores (n=309; 1.5 mm in diameter) were sampled from tumour-dense regions of archival formalin-fixed paraffin-embedded (FFPE) tissue blocks and assembled into tissue microarrays. These were subsequently profiled using the 10x Genomics Xenium platform for spatially resolved single-cell transcriptomics, employing a custom 344-gene panel designed to capture known transcriptional heterogeneity and cell type markers in gliomas (**Supplementary Table 2**). This approach yielded a dataset comprising 2.6 million nucleated cells, with a mean of 8,300 cells per core (**Figure 1D**). Cells were subsequently annotated using principles of single-cell RNA sequencing data analysis (**Methods**).

Of these nucleated cells, 82% were classified as tumour cells and 18% corresponded to 18 distinct non-neoplastic cell types constituting the TME (**Figure 1D; Extended Data Figure 2A**). Non-neoplastic cell types were annotated based on the expression of canonical marker genes (**Figure 1E, Extended Data Figure 2A, Supplementary Table 3**). This identified brain-resident populations, including astrocytes (*AQP4⁺/FGFR3⁺*), oligodendrocyte precursor cells/oligodendrocytes (OPC/OC; *PLP1⁺/OLIG1⁺*), and excitatory and inhibitory neurons (*STMN2⁺* and *GAD1⁺*, respectively), alongside an immune compartment comprising four myeloid subsets (*CD163⁺, P2RY12⁺, CLEC10A⁺,* and *RNASE1⁺* myeloid cells), neutrophils (*S100A9⁺*), lymphoid populations (CD4⁺ T cells, NK cells, CD8⁺ T cells, and cycling T cells, further distinguished by *KLRB1, GZMA, CD3G, CD8A, and TOP2A*), B cells (*CD19⁺*), plasma cells (*MZB1⁺*), and mast cells (*KIT⁺*). A separate vasculature compartment comprised vascular endothelial cells (*VWF⁺*) and perivascular fibroblasts (*COL3A1⁺*/*MYO1B⁺*). Tumour cells were distinguished by their expression of *EGFR* and *NES*. To distinguish astrocytes from tumour cells, a known challenge in glioma analysis ^7,10^, we used copy number variation (CNV) inference on an independent glioblastoma snRNA-seq dataset, which identified *FGFR3* as an astrocyte-specific marker (**Extended Data Figure 3**). Notably, the proportion of brain-resident non-malignant cells was low in the majority of samples, with only a few exceeding 50% (**Extended Data Figure 2B**). Most cases with low tumour cell content were low-grade astrocytomas.

Collectively, our data provide one of the largest and most comprehensive spatially resolved single-cell characterisations of adult gliomas to date, resolving both tumour and microenvironmental heterogeneity across the full spectrum of disease entities and grades.

**Figure 1:**
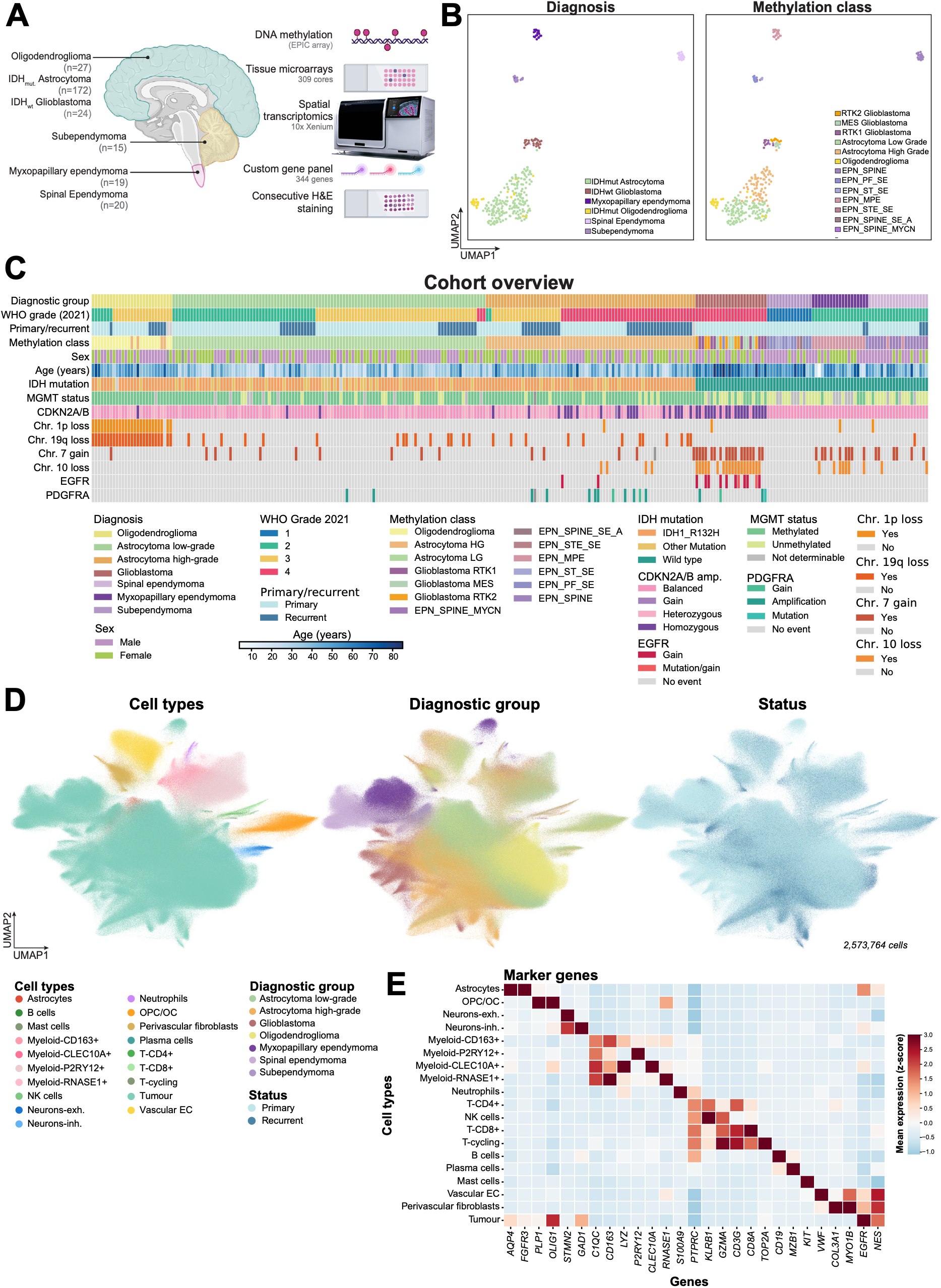
Spatial transcriptomics profiling of adult gliomas. **(A)** Overview of experimental workflow. Schematic illustrating the anatomical origins of CNS tumour samples. Tissues from various CNS regions (hemisphere, cerebellum, spinal cord) were processed using tissue microarrays (309 tumour cores, 282 sample specimens, 275 patients) and subjected to spatial transcriptomics (10x Xenium) with a custom 344-gene panel, followed by consecutive H&E staining. Created in BioRender. Mathioudaki, A. (2026) https://BioRender.com/q3gozpo. **(B)** UMAP of adult gliomas samples profiled using DNA methylation and coloured by diagnosis and DNA methylation class. **(C)** Clinical and molecular overview of the cohort. Each column represents one patient at one timepoint (primary or recurrent tumour; 280 specimens from 275 patients), grouped by diagnosis. Where several specimens from the same patient and primary/recurrent status were profiled, their annotations were combined. **(D)** UMAP of 2,573,764 cells, coloured by major cell types including tumour cells, OPC/OC, neurons, vasculature (endothelial and perivascular cells), myeloid cells, and lymphocytes. **(E)** Heatmap depicting scaled mean expression of marker genes. Abbreviations: CNS: central nervous system; H&E: hematoxylin and eosin; UMAP: Uniform manifold approximation and projection; OPC/OC: Oligodendrocyte progenitors/oligodendrocytes; EPN: Ependymoma.

### The recurrent tumour states of adult gliomas

While tumour cell states have been described within individual glioma subtypes in previous studies^10^, their comparative characterisation across glioma entities remains limited. To systematically address this, we fit a linearly decoded variational autoencoder (LDVAE) ^24,25^ model to the expression profiles of the tumour cells. This identified nine malignant cell states that recur across our cohort (**Figure 2A, B; Methods**).

This analysis recovered canonical tumour states previously described in glioblastoma, such as *AQP4*⁺ astrocyte-like (AC-like), *OLIG1*⁺ oligodendrocyte-progenitor-like (OPC-like), and SOX11^+^ neural-progenitor-like (NPC-like) states, as well as mesenchymal-like (*CD44*⁺ MES1 and *VEGFA*⁺ MES2) tumour cell states (**Figure 2B; Supplementary Table 4**). Notably, we resolved the OPC-like population into two substates, OPC-like I and II, with OPC-like I characterised by higher *PLP1* expression (**Figure 2B**). PLP1 encodes a protein of CNS myelin^26^. We further identified a proliferating state (*TOP2A*⁺, *MKI67*⁺), a stressed state (*HSPA1A⁺*), and a tumour state expressing ciliary markers (*KLHDC8A*⁺, *FANK1*⁺) enriched in ependymal tumours hereafter termed ependymal (EPN)-like tumour cells (**Figure 2A, B**). Genes defining different tumour states showed low co-expression within individual tumour cells (**Figure 2C**). The genes defining each state were consistent across diagnoses, allowing direct comparison of cell-state composition across glioma entities (**Extended Data Figure 4)**. Comparing tumour cell state abundance across diagnoses, we found that some states were diagnosis-restricted – OPC-like I, for instance, was largely specific to oligodendroglioma – while others were broadly shared across entities, albeit at varying frequencies (**Figure 2D**). Proliferating and AC-like tumour states were the most recurrent states, suggesting that these represent universal glioma programs. The occurrence of an AC-like tumour state in various tumour entities is in line with previous reports identifying astrocytic-like transcriptional programs in paediatric ependymomas ^22^. MES1/2-like states were most prevalent in high-grade astrocytoma and glioblastoma, occurring at lower frequency in other diagnoses except myxopapillary ependymoma (**Figure 2D**). In contrast, OPC-like and NPC-like states were largely restricted to diffuse gliomas, whereas the stressed and ependymal-like states were mostly found in ependymomas. Ependymal-like cells were also detected in glioblastoma, albeit at lower frequencies, consistent with studies reporting a subset of tumour cells exhibiting cilia-associated features ^27^. These findings reveal substantial differences in tumour state diversity across diagnoses, with glioblastoma exhibiting particularly high heterogeneity. At the same time, they highlight tumour state recurrence across genetically and epigenetically distinct glioma entities and provide new insights into less extensively characterised tumours, such as spinal ependymomas, which were predominantly composed of AC-like and ependymal-like states (**Figure 2D**).

Incorporating spatial information, we observed that these tumour cell states tend to occupy distinct, non-overlapping regions (**Figure 2E**). This spatial segregation suggests that tumour cell states are spatially organised rather than randomly intermixed within tumours.

**Figure 2:**
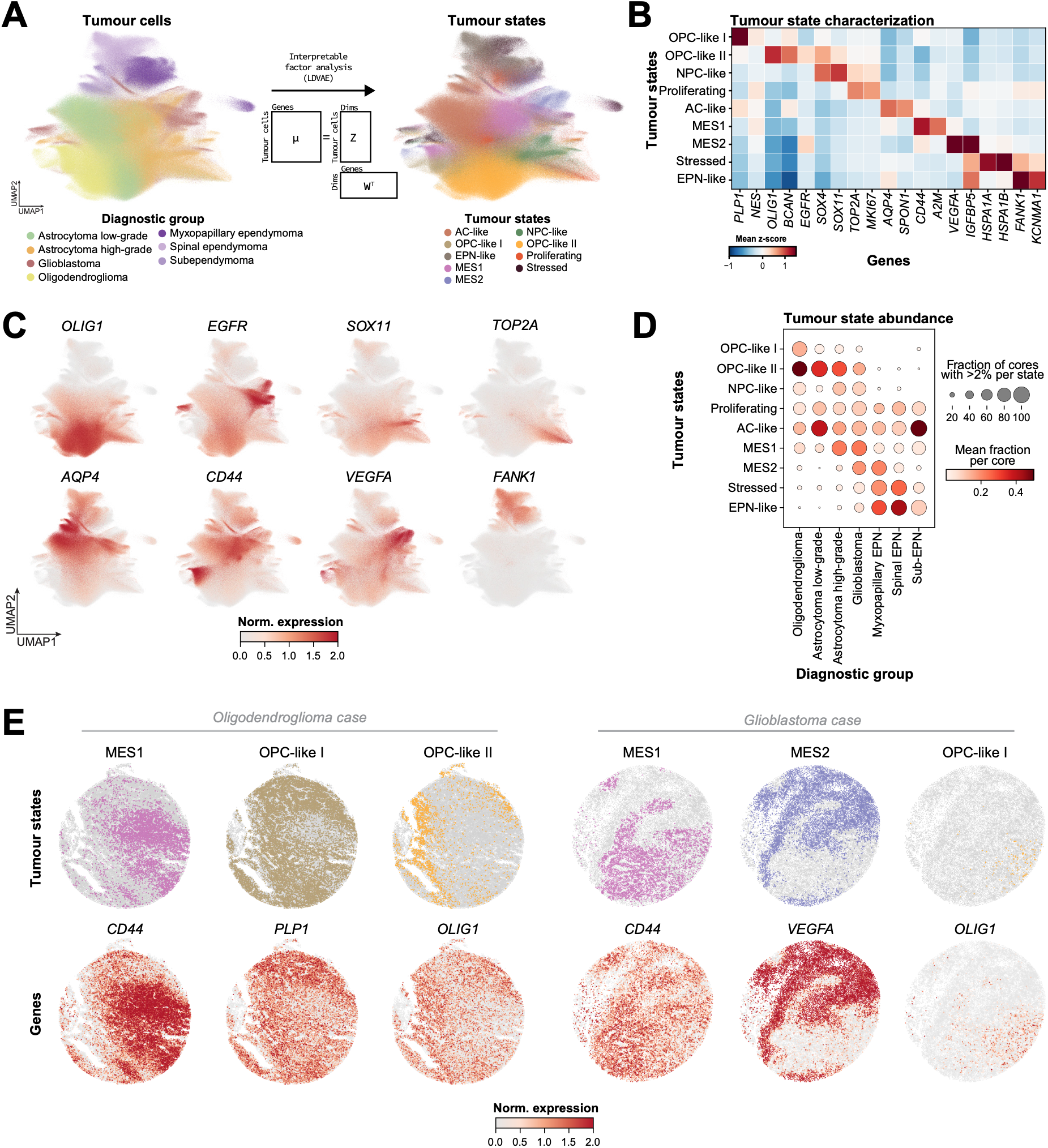
Characterisation of recurrent malignant cell states in adult gliomas. **(A)** UMAP embeddings of tumour cells coloured by tumour state (left) and by glioma entity (right). Tumour states were assigned using LDVAE. **(B)** Heatmap showing expression of key marker genes defining each tumour state across all tumour cells. **(C)** UMAP embeddings showing the log-normalised expression of marker genes. **(D)** Dot plot showing the fraction of each tumour state per core, stratified by glioma entity. Dot colour represents the mean fraction per core; dot size indicates the proportion of samples containing that neighbourhood. Only cores with >1% abundance per cell type were considered. **(E)** Representative spatial transcriptomics plots from an oligodendroglioma case (left) and a glioblastoma case (right), showing the spatial distribution of tumour cell states (top) and normalised expression of the corresponding state-defining marker genes (bottom). Abbreviations: LDVAE: linearly decoded variational autoencoders; UMAP: Uniform manifold approximation and projection; OPC: Oligodendrocyte progenitor; AC-like: Astrocyte-like; EPN: Ependymal/ependymoma.

### Glioma tumour states occupy distinct spatial neighbourhoods characterised by a unique TME

Building on the observation that tumour states exhibit spatial segregation rather than being uniformly intermixed, we next sought to characterise their spatial organisation and associated microenvironment. To do so, we defined spatial neighbourhoods as local environments characterised by the composition of neighbouring cells, using Delaunay-triangulation graphs constructed from cell coordinates (**Figure 3A, Methods**). In total, we identified 11 spatial neighbourhoods with distinct compositions of malignant and non-malignant cells (**Figure 3B, C**).

Most neighbourhoods were dominated by tumour cells and were distinguished primarily by the relative abundance of specific tumour states (**Figure 3C, D; Extended Data Figure 5A**). Among these, MES1-like cells predominantly occupied the gliosis neighbourhood ^12^ and MES2 the hypoxia neighbourhood ^28^. In contrast, one neighbourhood was characterised by a low tumour burden and the presence of perivascular and endothelial cells, resembling vasculature. A separate neighbourhood comprised tumour cells alongside brain-resident cell types, including oligodendrocytes, which we termed the tumour–brain resident interface (TBI) neighbourhood (**Figure 3C, D**). Overall, the cellular composition of these neighbourhoods was largely conserved across diagnoses (**Extended Data Figure 5B**).

Beyond their tumour cell state composition, neighbourhoods also differed in their relative abundance of brain-resident and peripheral immune cells (**Figure 3D**). The association of specific tumour states with immune-enriched neighbourhoods may indicate that some tumour phenotypes may be shaped or reinforced by microenvironmental cues. Specifically, MES1-and MES2-like states were associated with neighbourhoods enriched for peripheral immune cells such as CD163⁺ myeloid cells, consistent with previous reports linking mesenchymal and hypoxic states to myeloid-rich microenvironments (**Figure 3D**) ^12,28^. In contrast, the TBI, OPC-like, NPC-like, and AC-like neighbourhoods were enriched for brain-resident cells. These observations demonstrate that tumour states are spatially organised within the tissue microarchitecture rather than uniformly distributed throughout the tumour, with distinct states associated with specific microenvironmental cell populations, suggesting that tumour cell phenotypes may be influenced by local cellular interactions.

Across tumour diagnoses, neighbourhoods exhibited distinct distribution patterns, with some being entity-specific and others shared across histological subtypes (**Figure 3E, F**). In particular, the AC-like and proliferating neighbourhoods appear in all entities, including ependymomas, whereas other neighbourhoods were predominantly observed in individual entities, such as the OPC-like I neighbourhood in oligodendroglioma and the EPN-like neighbourhood in ependymomas. Within astrocytomas, DNA methylation profiling distinguishes two major subclasses, corresponding to lower- and higher-grade tumours ^2^. AC-like neighbourhoods were more abundant in low-grade astrocytomas, whereas hypoxia neighbourhoods were mostly observed in high-grade astrocytomas, corresponding largely to WHO grades 3 and 4 (**Figure 3F**). Interestingly, hypoxia neighbourhoods were also detected in myxopapillary ependymoma, a CNS WHO grade 2 tumour, indicating that these spatial programs are not restricted to high-grade tumours.

We next examined how these neighbourhoods relate to each other. This analysis revealed higher-level spatial layering among neighbourhoods: gliosis and hypoxia neighbourhoods frequently colocalised, NPC-like states are often adjacent to OPC-like regions, and AC-like cells were often positioned near OPC-like II and gliosis neighbourhoods (**Figure 3G, H**). The spatial proximity between AC-like and gliosis states may reflect a gradual transcriptional shift from astrocytic to mesenchymal programs, consistent with prior reports of reactive AC transitioning to MES states under proinflammatory stimuli ^15,29^. This spatial association was observed across multiple diagnoses, with particularly pronounced patterns in ependymal tumours and higher-grade gliomas (**Figure 3G, H; Extended Data Figure 6**).

Collectively, our data reveal that glioma tumour states occupy distinct spatial neighbourhoods characterised by specific malignant and non-malignant cell populations, highlighting how shared cellular programs are organised within distinct tissue microenvironments across entities.

**Figure 3:**
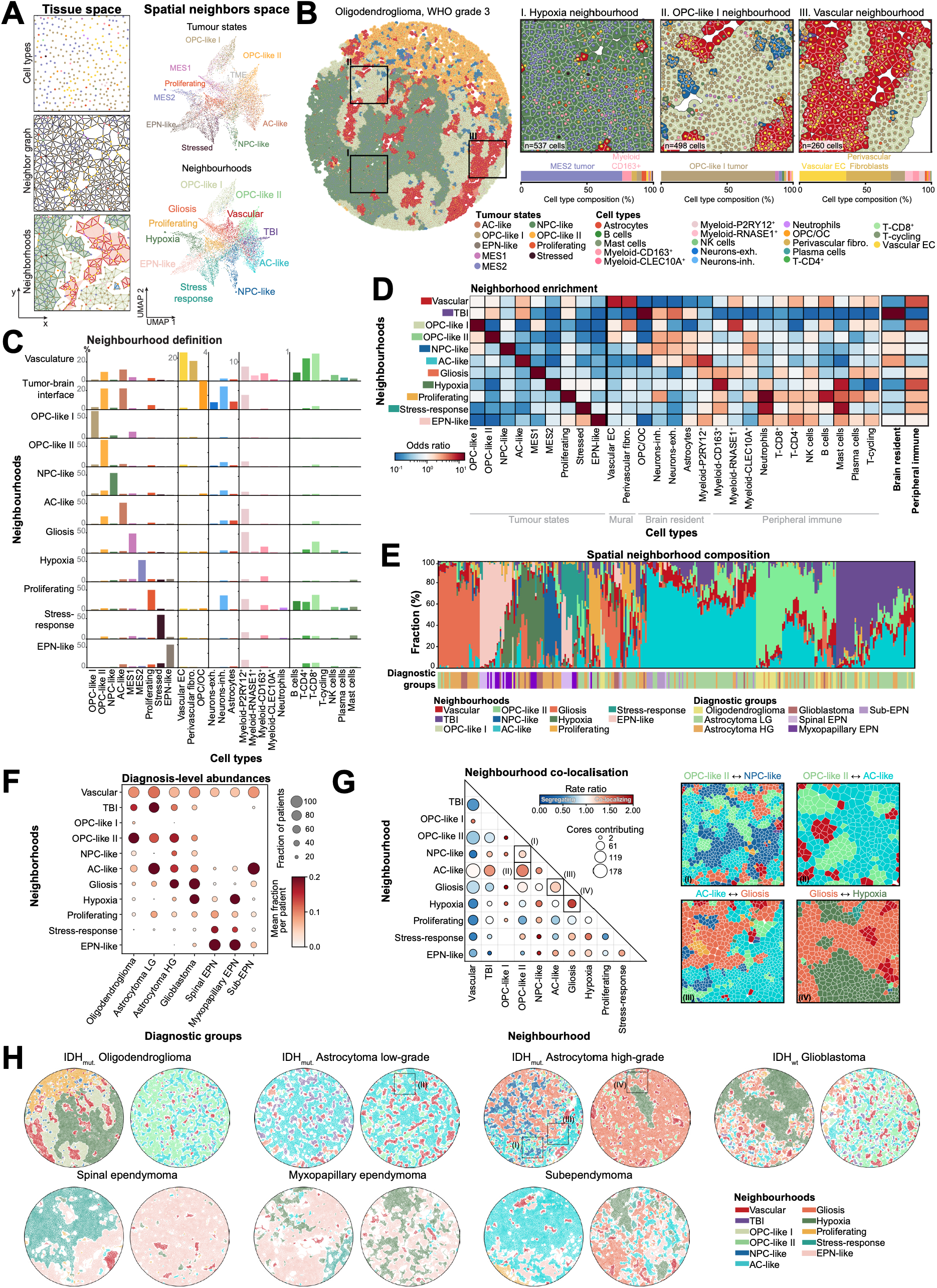
Microanatomical tissue organisation in adult-type gliomas. **(A)** (left) Schematic overview of the spatial neighbourhood analysis workflow, illustrating the construction of Delaunay triangulation graphs from spatial cell coordinates and the identification of spatial neighbourhoods. (right) UMAP representation based on spatial neighbourhood composition, coloured by the central tumour cell state (top) and assigned spatial neighbourhoods (bottom). **(B)** Representative case showing the spatial distribution of neighbourhoods, with magnified views of neighbourhoods. Corresponding cell type compositions are shown as bar plots below each region. **(C)** Cellular composition of the 11 identified spatial neighbourhoods, shown as the fraction of each cell type within each neighbourhood. **(D)** tumour-state (left) and tumour microenvironment (right) enrichment within spatial neighbourhoods. Enrichment is expressed as the odds ratio relative to the overall cellular composition of the dataset. **(E)** Neighbourhood composition per sample. **(F)** Dot plot showing the mean fraction of neighbourhoods per entity. Dot colour represents the mean fraction per core; dot size indicates the proportion of samples containing that neighbourhood. Only cores with >1% abundance per cell type were considered. **(G)** Left: co-localisation matrix of neighbourhood–neighbourhood contacts across entities. Dot colour indicates the Poisson rate ratio of observed versus permutation-expected contacts between spatially contiguous neighbourhood regions (>1, co-localisation; <1, segregation); dot size indicates the number of cores in which the neighbourhood pair was assessable. Right: Representative examples of co-localised neighbourhood pairs. **(H)** Representative tissue cores illustrating the higher-order spatial organisation of neighbourhoods in adult gliomas. Squares indicate regions highlighted in G. Abbreviations: UMAP: Uniform manifold approximation and projection; OPC-like: Oligodendrocyte progenitor-like; AC-like: Astrocyte-like; EPN: Ependymal/ependymoma.

### Spatial neighbourhood composition associates with clinical parameters in adult gliomas

The diagnosis of adult gliomas integrates histopathological features, DNA methylation profiles, and genetic alterations to establish tumour type and grade (**Figure 4A**). We therefore asked whether the spatial neighbourhood composition of tumours was associated with these established clinicopathological and molecular characteristics.

We first investigated associations between recurrent genetic alterations and spatial neighbourhood composition (**Figure 4B; Extended Data Figure 7; Supplementary Table 5**). Given the central role of IDH mutation status in glioma classification, association analyses were performed separately for IDH-wildtype and IDH-mutant gliomas.

Within IDH-mutant gliomas, the 1p/19q codeletion, a defining molecular feature of oligodendroglioma, was strongly associated with increased OPC-like neighbourhood abundance and reduced AC-like and gliosis neighbourhood abundance. Beyond this diagnosis-defining alteration, *CDKN2A/B* homozygous deletion was associated with differences in several spatial neighbourhoods, including increased representation of NPC-like, stressed, and ependymal-like neighbourhoods (**Figure 4B**). These associations are particularly relevant given the established role of *CDKN2A/B* homozygous deletion in the grading of IDH-mutant astrocytomas ^30^.

In IDH-wildtype glioblastoma, fewer significant associations were detected. *EGFR* alterations were associated with differences in EPN-like neighbourhood abundance, corresponding to a tumour cell population with cilia-associated features, whereas *PDGFRA* amplification was associated with reduced gliosis neighbourhood abundance (**Figures 2B, 4B**). Previous studies have linked *PDGFRA* amplification to increased OPC-like tumour states ^27^, a trend also observed in our data, although it did not reach statistical significance. These findings are compatible, as the relative abundance of OPC-like neighbourhoods may increase at the expense of gliosis neighbourhoods. Given the cohort size of 23 glioblastoma patients, additional genotype–neighbourhood associations may not have reached statistical significance. Since DNA methylation-based classification plays a central role in glioma diagnosis and prognosis, we next investigated whether spatial neighbourhood composition was associated with molecular subtypes. In glioblastoma, methylation subclasses exhibited distinct neighbourhood compositions, with Receptor Tyrosine Kinase subtype I (RTK I) tumours enriched for OPC-like II neighbourhoods, RTK II tumours for AC-like neighbourhoods, and MES tumours for gliosis neighbourhoods (**Figure 4C**).

Beyond molecular classification, we examined whether spatial neighbourhood composition was associated with WHO grade ^3^. Notably, WHO grade 4 IDH-mutant astrocytomas exhibited neighbourhood compositions more similar to glioblastomas, which are also WHO grade 4, than to lower-grade astrocytomas (**Figure 4D, Extended Data Figure 8A**). This suggests that spatial neighbourhood composition may reflect grade-associated tumour characteristics that extend across distinct molecular diagnoses.

Lastly, we sought to identify histopathological correlates of the transcriptionally defined neighbourhoods that could be detected using routinely generated hematoxylin and eosin (H&E)-stained sections. While necrotic regions, typically associated with MES2-tumour states and hypoxia, could be readily recognised by visual assessment, other neighbourhoods had less evident characteristics (**Figure 4E, Extended Data Figure 9**).

To quantify the association between neighbourhood prevalence and histopathology, we extracted embeddings from H&E images using the TITAN histopathology foundation model^31^ (**Figure 4E**). The histopathological feature space was only partially separated by diagnostic groups and IDH-mutant astrocytoma and IDH wildtype glioblastoma formed a continuum, which could be represented by four subclusters (**Figure 4F**). Spatial neighbourhood abundances varied noticeably across the histopathological feature space (**Figure 4E**). Within the histopathological continuum of astrocytoma and glioblastoma, neighbourhoods discriminated the set four subclusters more accurately than the diagnostic group alone (macro AUC 0.71 vs 0.62; **Figure 4H; Methods**). Cluster 1 contained tumours with a high abundance of the TBI and a depletion of other neighbourhoods, but was only weakly associated with diagnosis (**Figure 4I**). Similarly, cluster 3 was enriched for the gliosis neighbourhood. These findings link transcriptionally defined neighbourhood composition to H&E derived histomorphological patterns.

Collectively, our data demonstrate that spatial neighbourhoods capture clinically relevant tumour heterogeneity, associated with established molecular and clinicopathological features. Their association with histomorphology further raises the possibility of inferring molecular tissue organisation from routine histopathology, opening avenues for integrating spatial architecture into neuropathological characterisation of gliomas.

**Figure 4:**
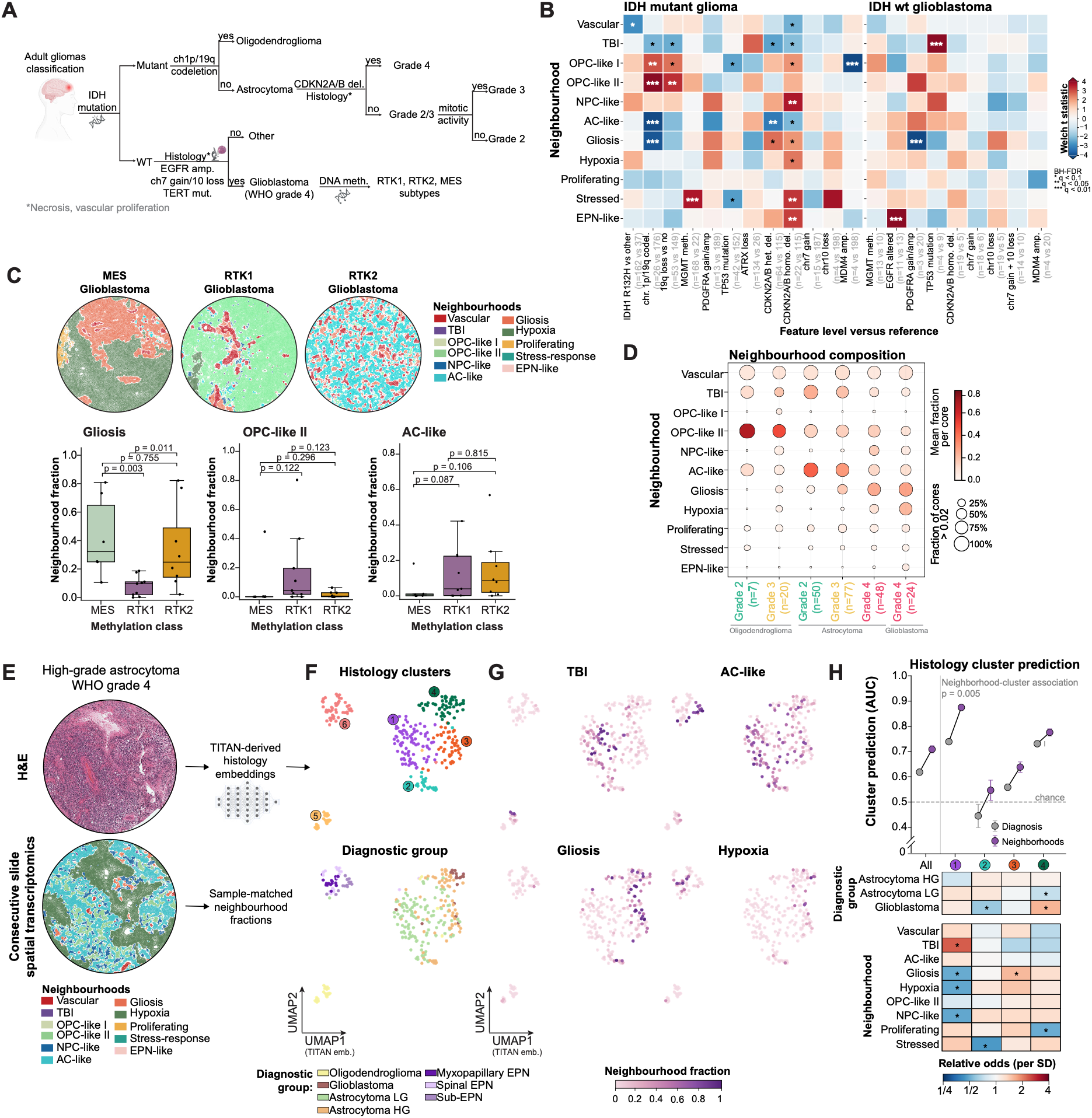
Linking spatially inferred tissue morphology with molecular and histopathological features. **(A)** Schematic overview of the parameters integrated into glioma diagnosis and classification. **(B)** Associations between genetic and molecular alterations and spatial neighbourhood composition in gliomas. Heatmaps show IDH-mutant gliomas (oligodendroglioma and astrocytoma; left) and IDH-wildtype glioblastoma (right). Each column compares tumours carrying the indicated alteration (test) with tumours without it (reference). Colour indicates the two-sided Welch t statistic computed on logit-transformed per-patient neighbourhood fractions. Asterisks indicate Benjamini–Hochberg-adjusted q-values, corrected across neighbourhoods within each comparison. Numbers in parentheses give the number of tumours (test vs reference). **(C)** (top) Representative glioblastoma tumour cores illustrating differences in neighbourhood architecture across methylation classes. (bottom) Neighbourhood abundance across glioblastoma DNA methylation classes. P values were calculated using two-sided Mann–Whitney U tests. **(D)** Dot plot showing the mean fraction of neighbourhoods across WHO grades and diagnoses. Dot colour represents the mean fraction per core; dot size indicates the proportion of samples containing that neighbourhood. Only cores with >1% abundance per neighbourhood were considered. **(E)** Representative example of a spatially annotated tumour core with its consecutive H&E-stained section, illustrating the workflow for H&E-derived analysis. **(F, G)** UMAP embeddings of H&E-derived morphological features extracted using TITAN. Points represent tumour cores coloured by histology cluster (F, top), diagnostic group (F, bottom) and by neighbourhood fractions (G). **(H)** Histology cluster prediction from diagnosis versus neighbourhood composition in astrocytomas (low- and high-grade) and glioblastoma. (Top) Cluster-wise and macro-averaged (All) AUC of multinomial logistic regression; mean ± SD of 20 repeats of patient-grouped 5-fold cross-validation. P value calculated from within-diagnosis permutation. (Bottom) Multinomial logistic regression including diagnosis and neighbourhoods; colour indicates the odds ratio per SD of each predictor. Asterisks indicate Benjamini–Hochberg-adjusted q-values from patient bootstrap, * q < 0.05. Abbreviations: RTK: Receptor Tyrosine Kinase; MES: Mesenchymal; EPN: Ependyma/Ependymoma; HG/LG: High/low grade; AUC: Area Under the Curve. Parts of this figure were created in BioRender. Mathioudaki, A. (2026). https://BioRender.com/cjtxx14; https://BioRender.com/1sb1zgu.

### AC-like and TBI neighbourhoods predict survival in IDH-mutant astrocytoma

Our histomorphological analysis revealed substantial heterogeneity among astrocytomas, which were distributed across multiple histological clusters and showed considerable overlap with glioblastomas (**Figure 4D, F**). At the same time, WHO grade provided limited prognostic separation between grade 2 and grade 3 IDH-mutant astrocytomas (**Extended Data Figure 8B**). Together, these observations prompted us to investigate whether neighbourhood composition could provide additional prognostic information beyond the established WHO grading. Given the broad representation of IDH-mutant astrocytomas in our cohort (n=175), spanning WHO grades 2–4, we focused subsequent analyses on this tumour type.

To assess the relationship between spatial organisation and clinical stratification, we stratified tumours based on their most prevalent neighbourhood (**Figure 5A**). This revealed a dichotomy in lower-grade IDH-mutant astrocytomas, with tumours primarily characterised by either the tumour-brain-resident interface (TBI) or the AC-like neighbourhood (**Figure 5B, C**). This separation became progressively less pronounced with grade and was largely absent in grade 4 tumours, where additional neighbourhoods, such as the gliosis neighbourhood, became more prevalent (**Figure 5C**).

To further dissect these neighbourhoods, we profiled an independent cohort of 18 low-grade IDH-mutant astrocytoma samples using same-slide high-plex spatial transcriptomics and multiplexed immunofluorescence (5,096 genes and 15 proteins; **Figure 5D; Extended Data Figure 10; Supplementary Tables 6, 7**). As both assays were performed on the same section, we were able to perform joint analysis and feature transfer across modalities (**Methods**). The resulting multimodal dataset is publicly accessible at https://gliomap.dkfz.de/ for interactive exploration. This validated our spatial annotations at the protein level and provided additional morphological insights, including vascular structures surrounded by astrocytic endfeet, consistent with blood-brain barrier structure (**Figure 5D**). Our analysis confirmed that AC-like neighbourhoods were enriched for brain resident cells such as microglia, but contained few oligodendrocytes, which were instead enriched in the TBI neighbourhood (**Figure 5E**). Accordingly, TBI neighbourhoods were characterised by tumour cells interspersed among oligodendrocytes, with enrichment of myelin associated marker MAG, suggesting that these tumour cells may reside in proximity to myelinated axons (**Figure 5E**).

Having characterised these neighbourhoods and their association with astrocytoma grade, we next asked whether spatial composition holds prognostic value independent of existing clinical and molecular parameters. Survival analysis and multivariate Cox proportional hazards modelling revealed that enrichment for the AC-like neighbourhood was associated with significantly longer overall survival (OS), while dominance of the TBI neighbourhood correlated with poorer prognosis (**Figure 5F, Extended Data Figure 11; Supplementary Table 8**). Similar trends were observed for progression-free survival (PFS) and in analyses stratified by primary and recurrent tumours, with AC-like enrichment showing consistent associations with favourable outcomes across analyses (**Extended Data Figure 11**).

We next sought to validate these prognostic associations in independent cohorts. Our study represents one of the first large-scale, single-cell-resolution spatial transcriptomic characterisations of gliomas, and comparable spatial datasets remain scarce. Transcriptional signatures have previously been shown to carry prognostic significance across tumour types^32^. Spatial transcriptomics extends this approach by enabling the derivation of signatures which capture combined transcriptional programs of multiple cell types within spatially defined neighbourhoods. We therefore derived transcriptional signatures from aggregated neighbourhood expression profiles and quantified their expression in two independent bulk RNA-seq cohorts of a total of 555 patients (**Methods; Extended Data Figure 12; Supplementary Tables 8, 9**) ^33,34^. Stratification of patients into tertiles based on neighbourhood signature scores revealed that higher AC-like scores were associated with superior OS, whereas higher TBI scores were associated with poorer outcomes (**Figure 5G**). These results indicate that the prognostic value of the AC-like and TBI neighbourhoods extends beyond our own cohort, supporting their potential as generalizable biomarkers of clinical outcome in IDH-mutant astrocytoma.

Together, our findings establish spatial architecture as a core determinant of glioma clinical behaviour, complementary to current molecular classification.

**Figure 5:**
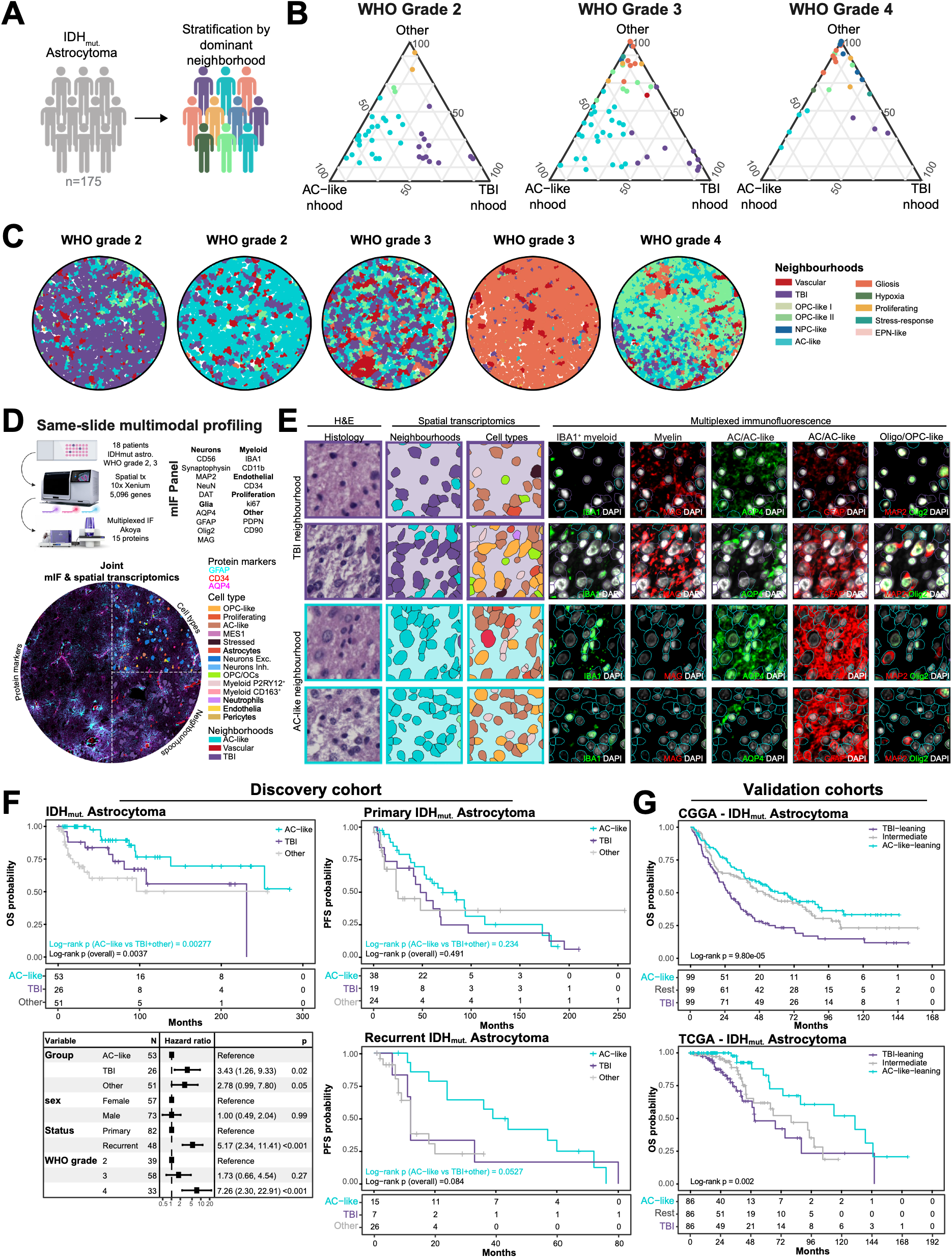
Neighbourhood landscape stratifies survival in IDH-mutant Astrocytoma. **(A)** Schematic illustrating patient stratification based on their most dominant spatial neighbourhood. **(B)** Ternary plots representing the relative proportions of TBI neighbourhood, AC-like neighbourhood, and other neighbourhoods in astrocytoma patients. Each point represents a patient sample, coloured by their most abundant neighbourhood. **(C)** Representative astrocytoma tumour cores illustrating differences in neighbourhood architecture across WHO grades. **(D)** Experimental workflow for same-slide spatial transcriptomics and multiplexed immunofluorescence (IF). **(E)** Joint H&E, protein and transcriptome profiling reveals distinguishing features of the TBI and AC-like neighbourhoods. **(F)** Kaplan–Meier curves for IDH-mutant astrocytomas in the discovery cohort, grouped by their most abundant spatial neighbourhood: AC-like, TBI, or other neighbourhoods (Other). Left, overall survival (OS) for all samples (n = 130 samples from 125 patients, including 5 with both primary and recurrent tumours). Forest plot: hazard ratios (HRs) with 95% confidence intervals and P-values from a multivariable Cox proportional hazards model. Right, progression free survival (PFS) shown separately for primary and recurrent tumours. Tick marks indicate censored observations; numbers at risk are shown below its plot. P-value: two-sided log-rank test across all three groups (overall) and for AC-like versus the other two groups combined (AC-like). **(G)** Kaplan–Meier curves of OS for IDH-mutant astrocytomas in two external cohorts, TCGA (n = 258 patients; 58 deaths) and CGGA (n = 297 patients; 192 deaths), grouped into tertiles of the balance score between the AC-like and TBI signatures (z(AC-like) − z(TBI), computed within each cohort). Numbers at risk are shown below the curves. P-value: two-sided log-rank test across the three tertiles. Abbreviations: TBI: tumour-brain resident interface; AC-like: astrocyte-like. Parts of this figure were created in BioRender. Mathioudaki, A. (2026). https://BioRender.com/k5gvfbg; https://BioRender.com/8lwgcu1.

## Discussion

Despite their distinct molecular and histopathological characteristics, adult-type glial tumours share therapeutic challenges, with many treatments applied across tumour types with variable efficacy ^1,19^. This overlap underscores the need to understand the commonalities among these tumours. By combining spatially resolved molecular profiling with histopathological analysis across IDH-wildtype and IDH-mutant gliomas and ependymomas, we show that tissue architecture is an integral component of glioma heterogeneity, encompassing both shared and entity-specific patterns of spatial organisation.

Tissue architecture, as assessed through MRI and histopathology, has long been recognised as an important feature of glioma biology^3^. Our findings provide molecular and cellular correlates of this architecture, resolving 11 recurrent neighbourhoods that differ in their immune and glial landscapes and in the tumour states they comprise. Proliferating and AC-like neighbourhoods were present across gliomas, whereas the remaining neighbourhoods aligned with diagnosis, methylation class and WHO grade. For instance, hypoxia/MES2 neighbourhood is enriched in glioblastoma and high-grade astrocytoma but is also detected in myxopapillary ependymoma, a WHO grade 2 tumour, suggesting that hypoxia-related programs can arise independently of tumour grade. Although mesenchymal signatures are associated with poor radiation response and reduced survival in glioblastoma^35^, and hypoxia with poor prognosis across solid cancers^36,37^, its presence in myxopapillary ependymoma suggests that this spatial program is not restricted to high-grade tumours.

The relevance of spatial architecture extends beyond tumour classification to patient outcomes. In astrocytoma, AC-like neighbourhood enrichment is associated with longer survival, whereas tumour–brain interface (TBI) enrichment is linked to poorer outcomes. These associations highlight spatial architecture as a complementary source of prognostic information beyond WHO grade.

The AC-like neighbourhood is particularly notable for its recurrence across entities and proximity to multiple tumour states, placing it at the center of the tumour-state landscape. AC-like programs have also been described in pediatric ependymomas ^22^, and we extend this to adult ependymoma subtypes. This broad distribution is compatible with AC-like representing a shared feature of glioma biology, and it raises the question of whether additional factors, such as immune cell recruitment, drive its diversification into other tumour states. Beyond its broad occurrence, AC-like tumour state has been associated with favorable response to IDH-mutant inhibition ^38^. Together with our findings on improved survival, this suggests that AC-like programs may have broader clinical relevance, although their prognostic significance beyond astrocytomas remains to be determined.

In contrast, the TBI neighbourhood was associated with poorer prognosis and comprised OPC-like and AC-like tumour cells interspersed among oligodendrocytes. Given the diffuse infiltrative growth of astrocytomas and the role of oligodendrocytes in axonal myelination, this organisation may reflect direct interactions between oligodendrocytes and tumour cells ^39^, or tumour cells positioned along axons ^40,41^. These associations may be related to the infiltrative growth pattern of astrocytomas, although direct tumour–axon interactions cannot be resolved through transcriptomic profiling alone.

Incorporating neighbourhood prevalence could refine existing classification systems. While spatial transcriptomics offers high resolution, its cost and complexity currently prevent routine clinical use, whereas H&E histopathology remains rapid, inexpensive and universally available. Advances in foundation models for digital pathology now enable extraction of high-dimensional morphological features from standard H&E slides ^42,43^. Consistent with this, we find parallel trends between spatial neighbourhoods and histopathology-derived features, a first step toward integrating molecularly informed tissue architecture into routine diagnostic workflows.

Our study has limitations. First, spatial sampling was based on tumour cores, which may not capture the full architecture of higher-grade tumours, where heterogeneity is greater ^16^. Second, the transcriptional signature of the AC-like tumour state closely resembles that of non-malignant astrocytes, so cell type assignments in these compartments may partly reflect both malignant and normal populations. Lastly, since the TBI neighbourhood comprises AC-like tumour, OPC-like tumour and oligodendrocytes, the signature extracted for validation in external cohorts reflects a combination of OPC-like and TBI signals. This may limit the attribution of the prognostic association to a specific neighbourhood, although both are associated with poorer outcomes.

Together, our findings show that glioma heterogeneity is organised in space: tumour states are not distributed uniformly but assemble into recurrent neighbourhoods, some shared and others specific to diagnosis and grade. This organisation adds an axis to classification that genetics and bulk profiling do not capture, and it links molecular programs to the tissue architecture that pathologists and radiologists already recognise. By scaling spatial profiling to hundreds of patients and linking it to routine histology, this work shows how high-throughput tissue mapping can move from describing glioma biology to informing glioma classification in the clinic.

## Data Availability

All processed and annotated spatial transcriptomics and snRNA-seq data objects have been deposited to Zenodo.
To adhere to local regulations, DNA methylation, H&E and clinical outcome data can be shared after completion of a data transfer agreement for noncommercial use. Requests should be made to the corresponding author(s), with responses expected within 14 days of receipt of request.

## Extended Data Figures

**Extended Data Figure 1:**
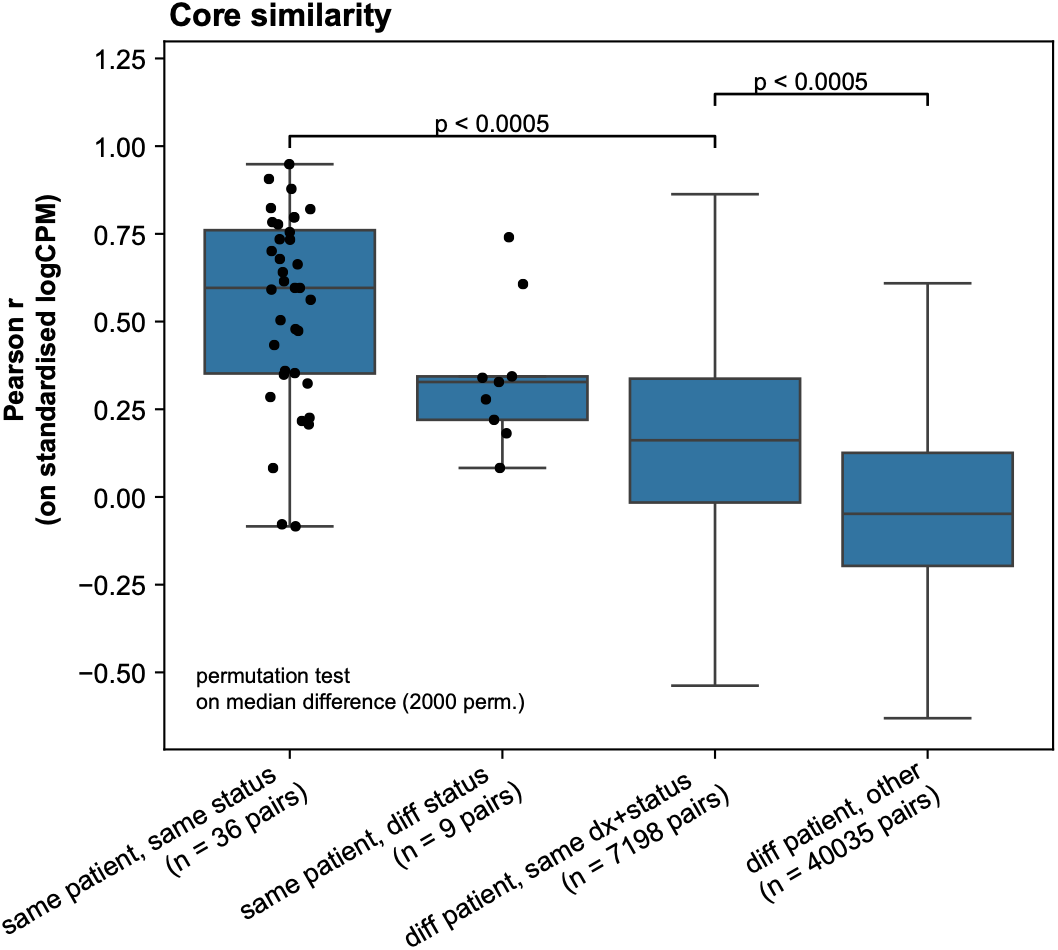
Expression profiles of TMA cores are more similar within than between patients. Pearson correlation of gene-wise standardised pseudobulk expression (log CPM) between pairs of TMA cores (308 cores from 275 patients), grouped into four categories: cores from the same patient and the same primary/recurrent status (n = 36 pairs); cores from the same patient taken at primary diagnosis and recurrence (n = 9); cores from different patients with the same diagnosis and status (n = 7,198); all other pairs (n = 40,035). Black dots show individual same-patient pairs. Boxes show the median and interquartile range, and whiskers extend to 1.5×the interquartile range; outliers are not shown. P-values are from one-sided permutation tests on the difference in median correlation (2,000 permutations). To account for non-independence of pairs, patient labels were shuffled between cores within each diagnosis–status stratum, or diagnosis–status labels were shuffled between groups of cores from the same patient and status. 24 patients were represented by more than one core: 18 by two cores, three by three and three by four. Cores from the same patient and status formed 22 replicate groups comprising 50 cores, and 5 patients contributed both primary and recurrent tumour tissue.

**Extended Data Figure 2:**
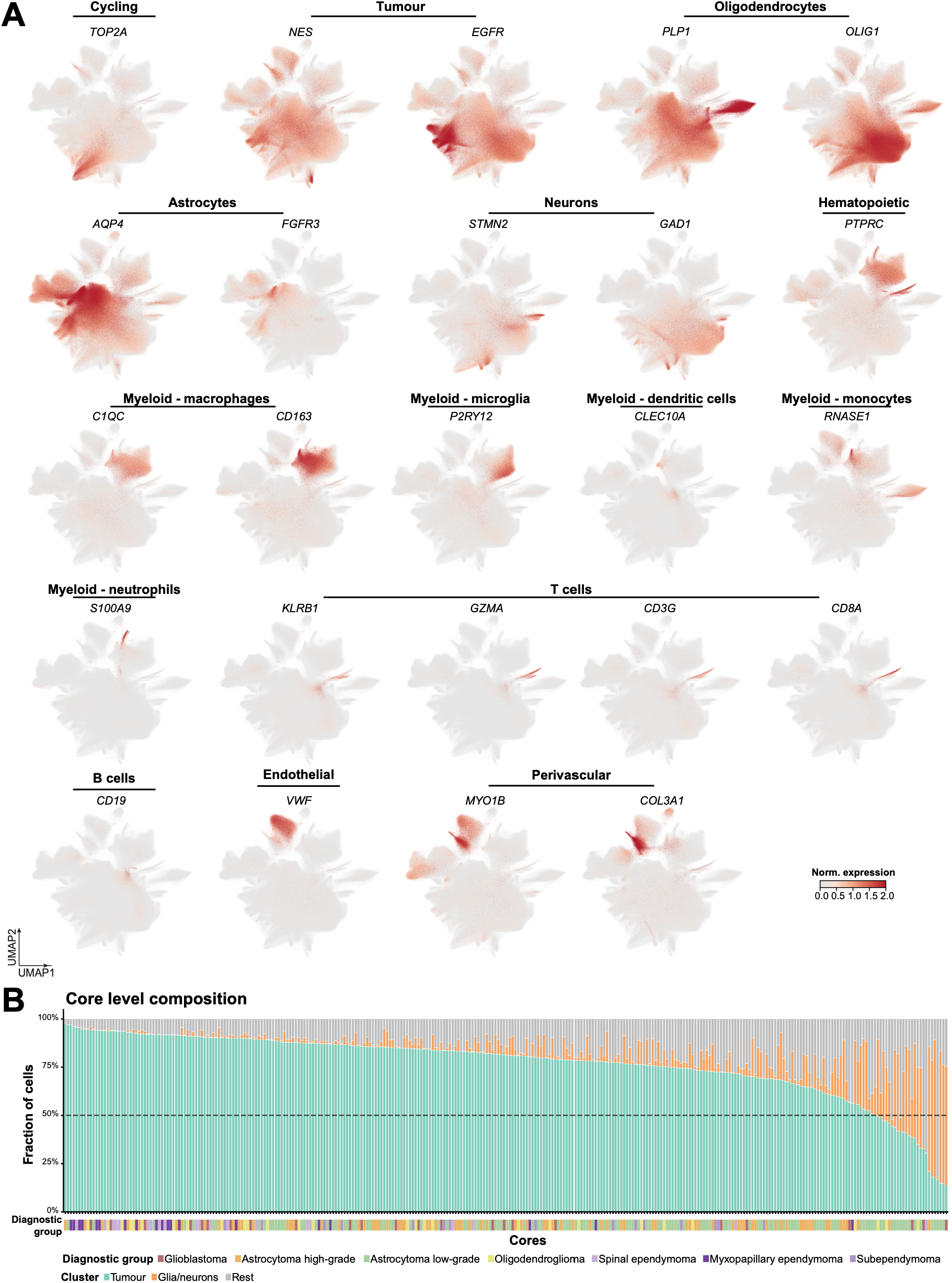
Spatial transcriptomics sample composition and marker genes. **(A)** Uniform Manifold Approximation and Projection (UMAP) embedding of all cells profiled by spatial transcriptomics, coloured by normalised expression of the marker genes used for cell-type. **(B)** Cell-type composition of each core, shown as the fraction of cells assigned to tumour, glia/neuron, and remaining clusters (stacked bars, one per core), ordered by decreasing tumour fraction. Diagnosis of each core is indicated below the plot.

**Extended Data Figure 3:**
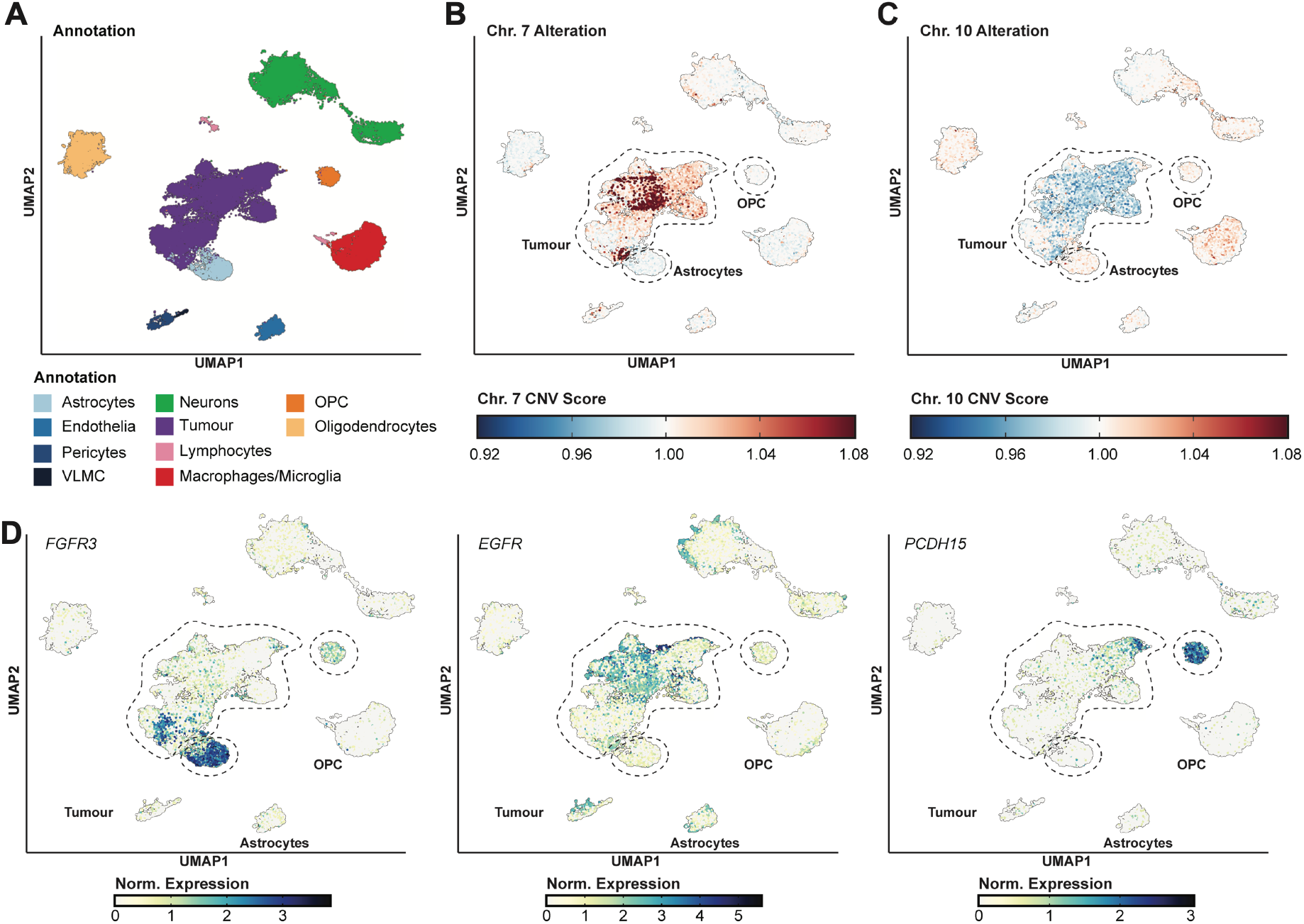
Identification of markers for confident tumour annotation. **(A)** UMAP representation of the merged snRNA-seq dataset, coloured by cell-type annotation. **(B, C)** UMAP plots coloured by inferred copy-number variation (CNV) scores for chromosome 7 (B) and chromosome 10 (C); tumour, OPC, and astrocyte clusters are outlined. **(D)** UMAP plots showing normalised expression of *FGFR3*, *EGFR*, and *PCDH15*, with the same clusters outlined.

**Extended Data Figure 4:**
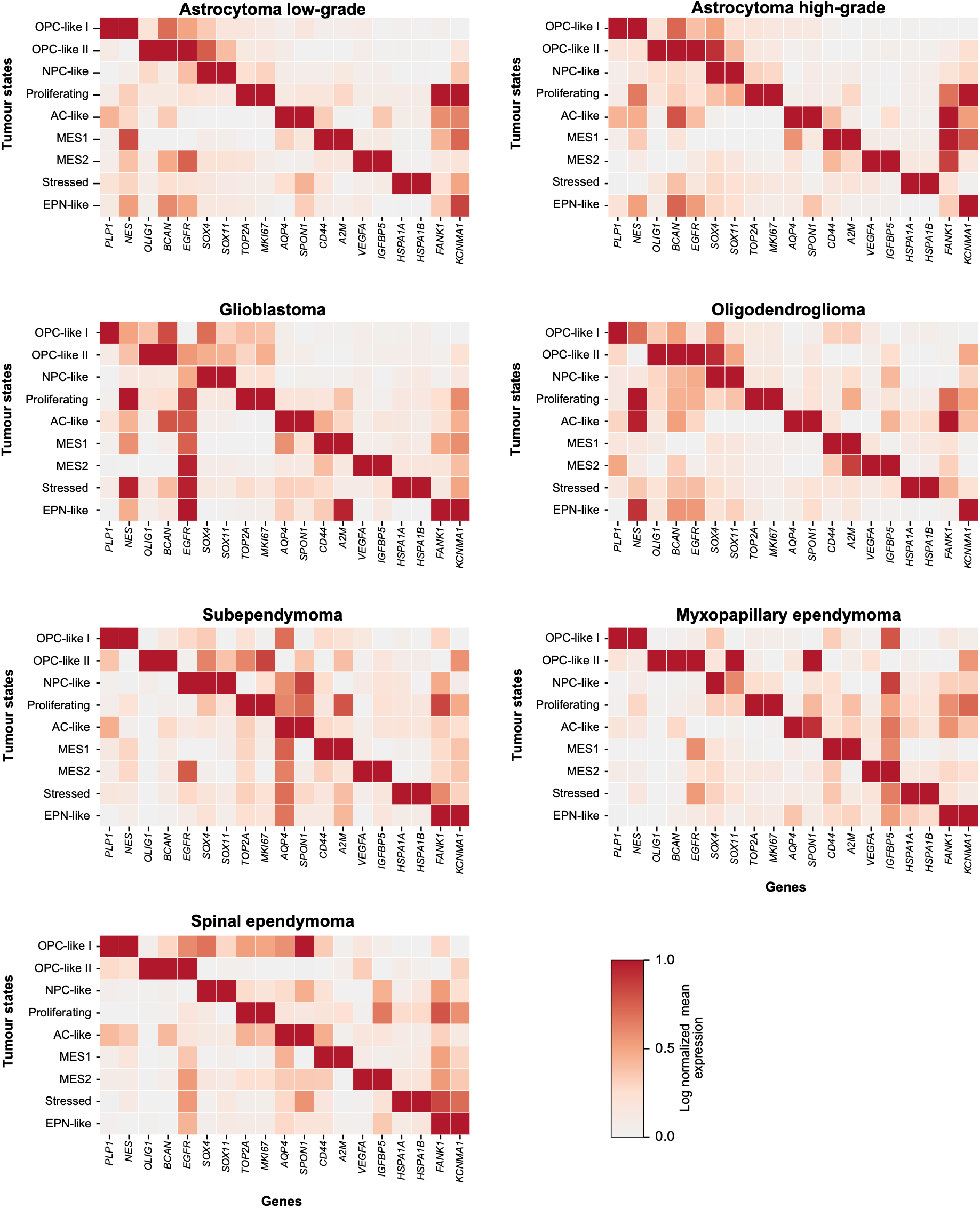
Tumour factor definition consistency across diagnoses. Heatmaps show the mean log-normalised expression of selected marker genes (columns) in tumour cells assigned to each of the nine tumour programs (rows), calculated separately for each diagnosis (n=1,629,748 tumour cells). Each cell was assigned to the tumour program with its highest latent factor weight. Numbers in the tiles show the mean expression. Colours are min–max scaled per gene within each pane.

**Extended Data Figure 5:**
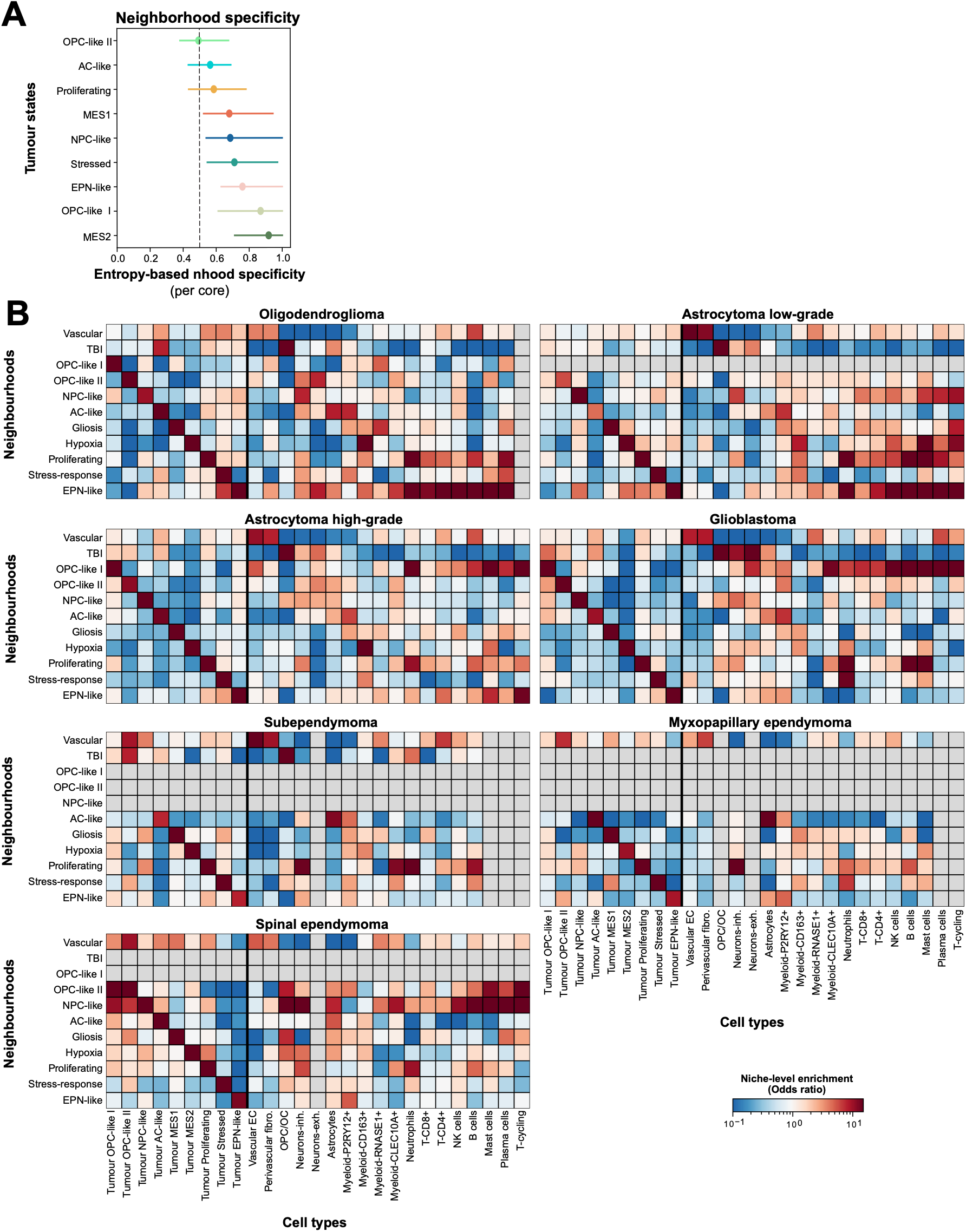
neighbourhood composition across diagnoses. **(A)** Spatial neighbourhood specificity of malignant tumour states, quantified using entropy. **(B)** tumour-state (left) and tumour microenvironment (right) enrichment within spatial neighbourhoods, shown separately for each diagnosis. Enrichment is expressed as the odds ratio relative to the overall cellular composition of that diagnosis. Grey cells indicate niches with fewer than 30 cells or cell types with fewer than 20 cells in the respective diagnosis.

**Extended Data Figure 6:**
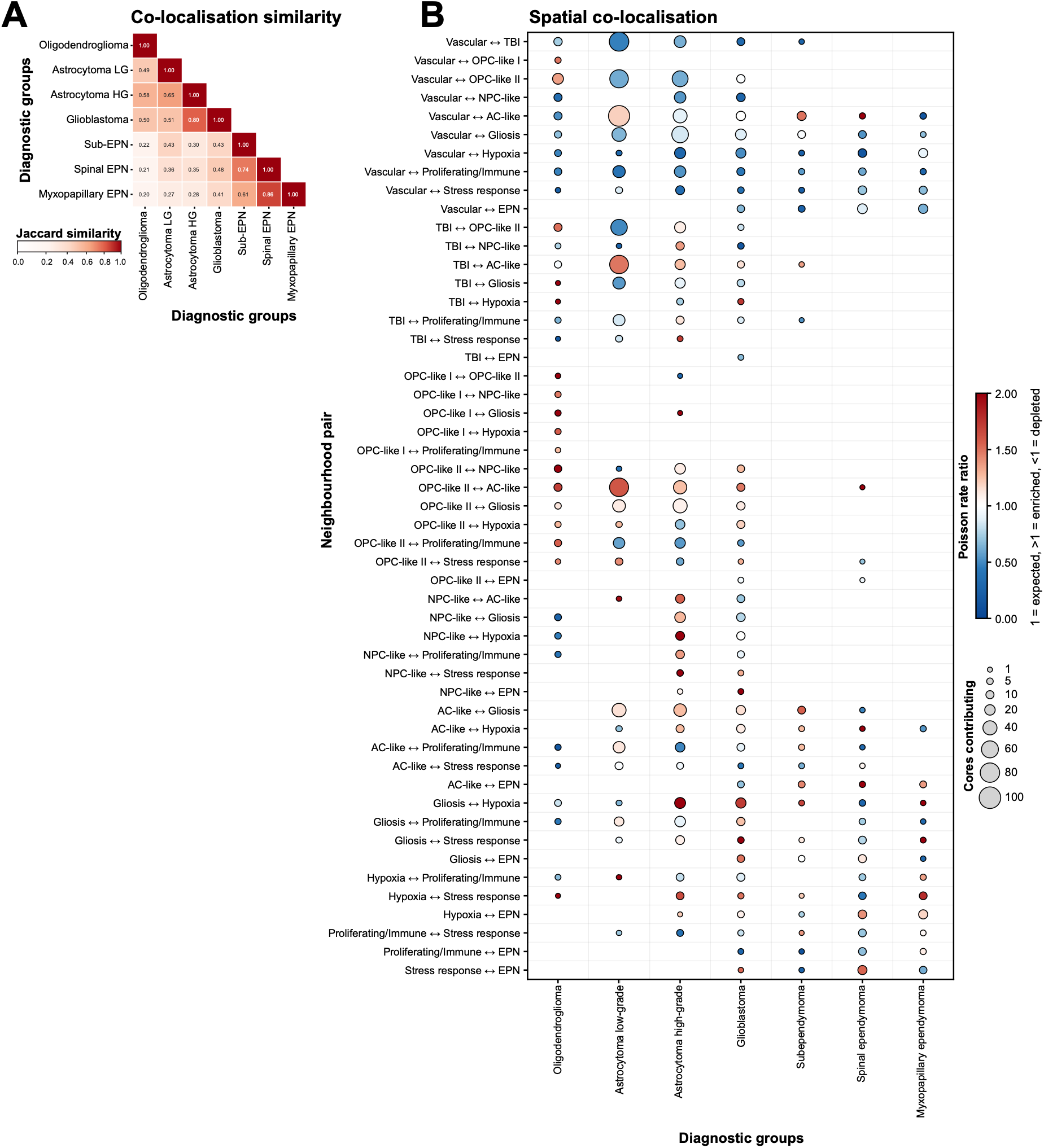
neighbourhood colocalisation. **(A)** Similarity of neighbourhood-pair repertoires across entities, quantified as the Jaccard index of neighbourhood pairs assessable in ≥2 cores per entity. **(B)** Diagnosis-specific neighbourhood–neighbourhood spatial interactions. Each dot represents one neighbourhood pair (rows) in one diagnostic entity (columns). Dot colour indicates the Poisson rate ratio of observed versus permutation-expected contacts between spatially contiguous neighbourhood regions, estimated separately for each entity (1, as expected; >1, enrichment; <1, depletion). Dot size indicates the number of cores in which the neighbourhood pair was assessable (non-zero expected contact count); sizes are capped at 100 cores. Only neighbourhood pairs assessable in ≥2 cores of an entity are shown.

**Extended Data Figure 7:**
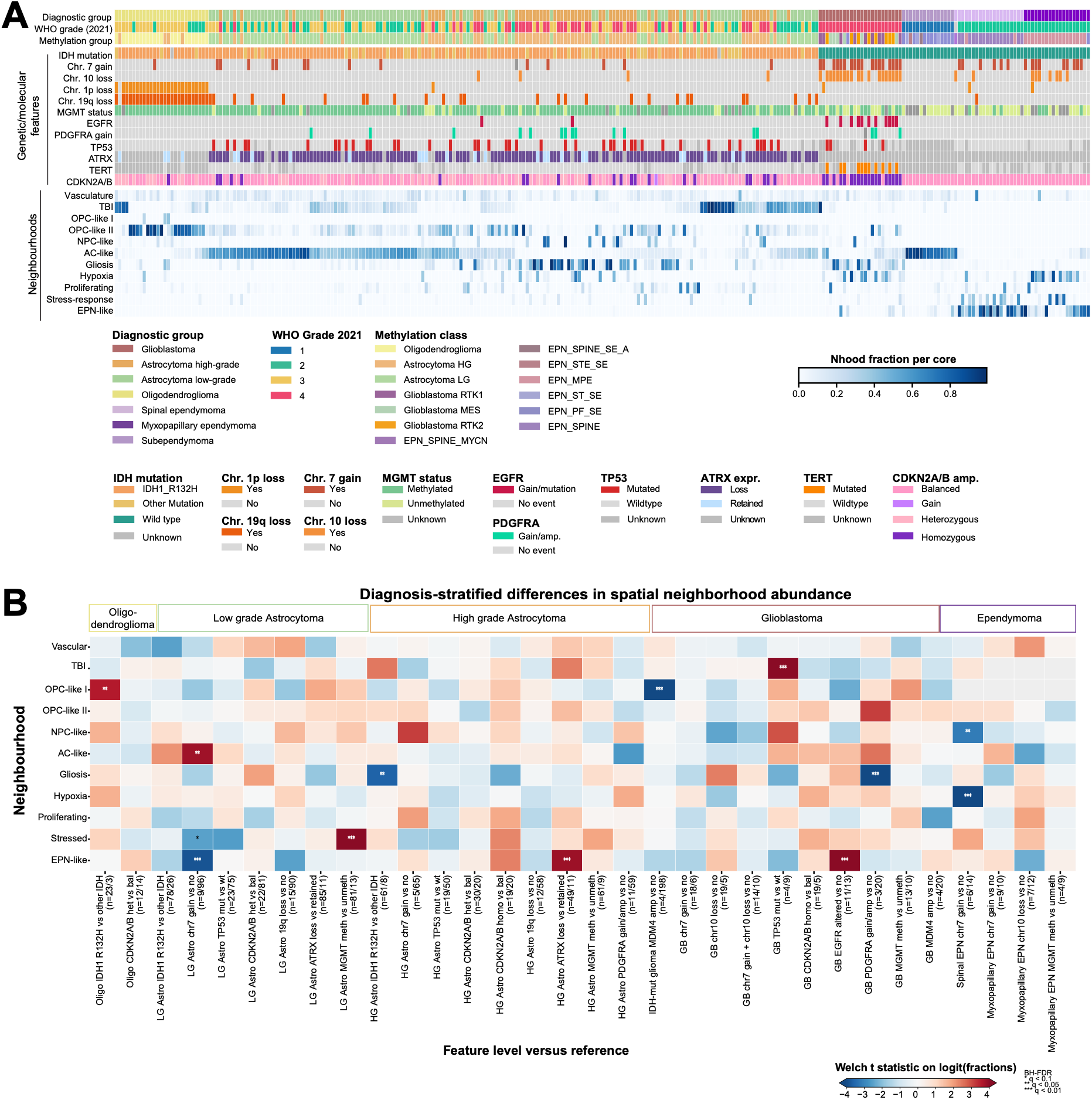
The impact of molecular and genetic events on neighbourhood abundance. **(A)** Overview of neighbourhood fraction per sample. **(B)** Diagnosis-stratified association of molecular and genetic alterations with spatial neighbourhood abundance. Contrasts were performed separately within diagnostic groups. Each column compares tumours with the indicated alteration against tumours without it within the same diagnosis. Colour shows the Welch t statistic computed on logit-transformed per-patient neighbourhood fractions. Asterisks show Benjamini–Hochberg-adjusted q-values, corrected across neighbourhoods within each contrast: * q < 0.1, ** q < 0.05, *** q < 0.01. Numbers in parentheses give the number of tumours in test versus reference. Contrasts with fewer than 3 tumours in either group were not tested and are not shown.

**Extended Data Figure 8:**
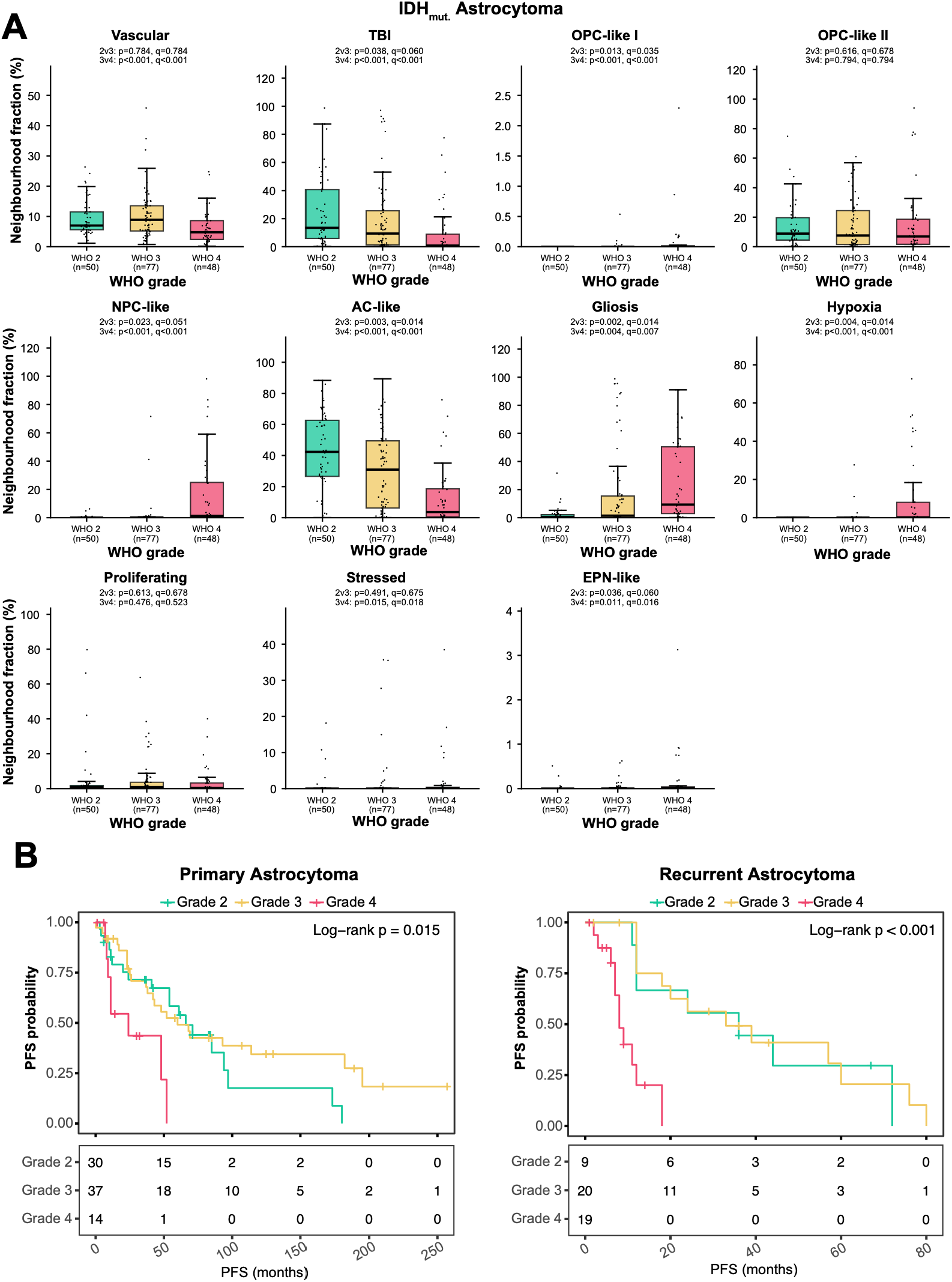
WHO grade 4 IDH-mutant astrocytomas are distinguished from lower-grade tumours by neighbourhood composition and shorter progression-free survival. (A) Fraction of each spatial neighbourhood per sample in IDH-mutant astrocytomas, stratified by WHO CNS grade. Each dot represents one sample. Boxes show the median and interquartile range; whiskers extend to the most extreme values within 1.5× the interquartile range. Pairwise comparisons between grade 2 and grade 3 (2v3) and between grade 3 and grade 4 (3v4) were performed using two-sided Mann–Whitney U tests. p, nominal p-value; q, Benjamini–Hochberg-adjusted p-value. (B) Kaplan–Meier estimates of progression-free survival (PFS) for patients with primary (left) and recurrent (right) astrocytoma, stratified by WHO CNS grade. Tick marks indicate censored observations. P-values were calculated using the log-rank test. Tables below each plot show the number of patients at risk at the indicated time points.

**Extended Data Figure 9:**
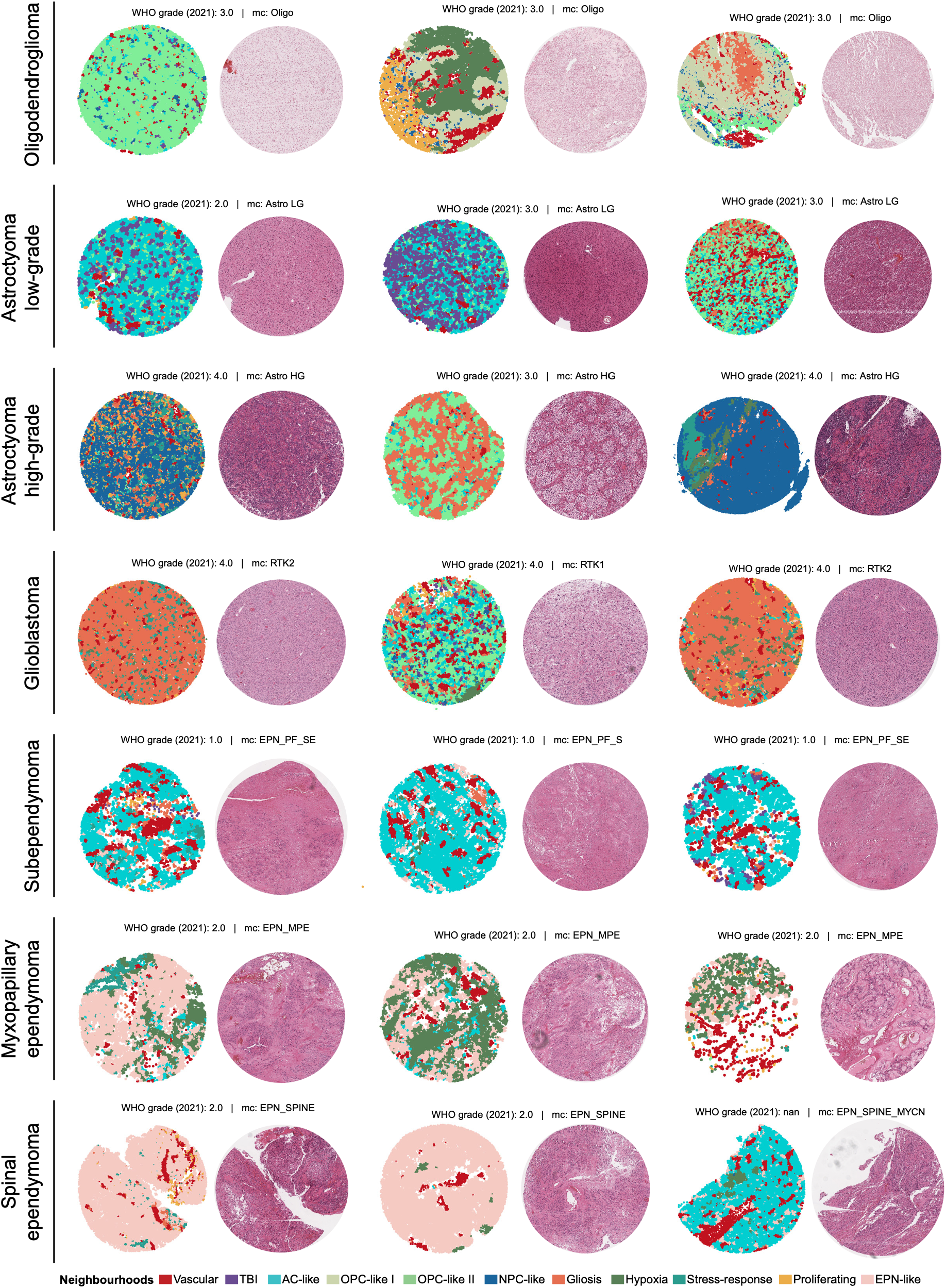
Spatial neighbourhood maps and matched H&E images of example cores across glioma entities. For each diagnostic group, three example TMA cores are shown. Each pair of panels shows the spatial map of the 11 spatial neighbourhoods (left; each cell is coloured by its neighbourhood assignment) and the corresponding H&E image of a consecutive section (right).

**Extended Data Figure 10:**
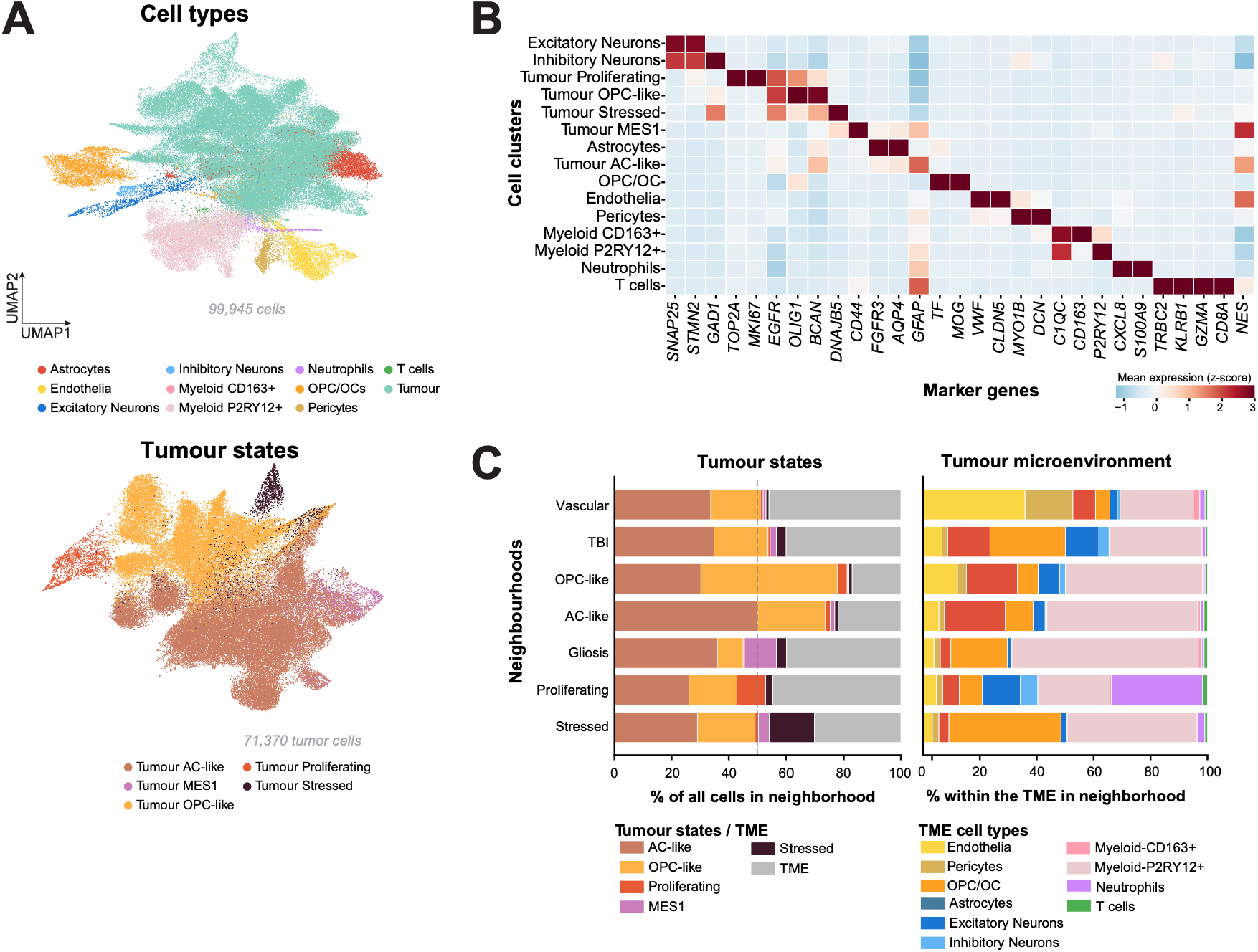
Same slide multimodal profiling of IDH-mutant astrocytoma. **(A)** UMAP embedding of 99,945 cells from 18 astrocytoma patients, coloured by major cell type (left) and tumour cell state (right). Samples were profiled with the Xenium spatial transcriptomics platform using a 5,096-gene panel. **(B)** Heatmap of scaled mean expression of marker genes across cell types and tumour states. **(C)** Neighbourhood composition of the same samples. Bars show the proportion of a given cell type within each neighbourhood.

**Extended Data Figure 11:**
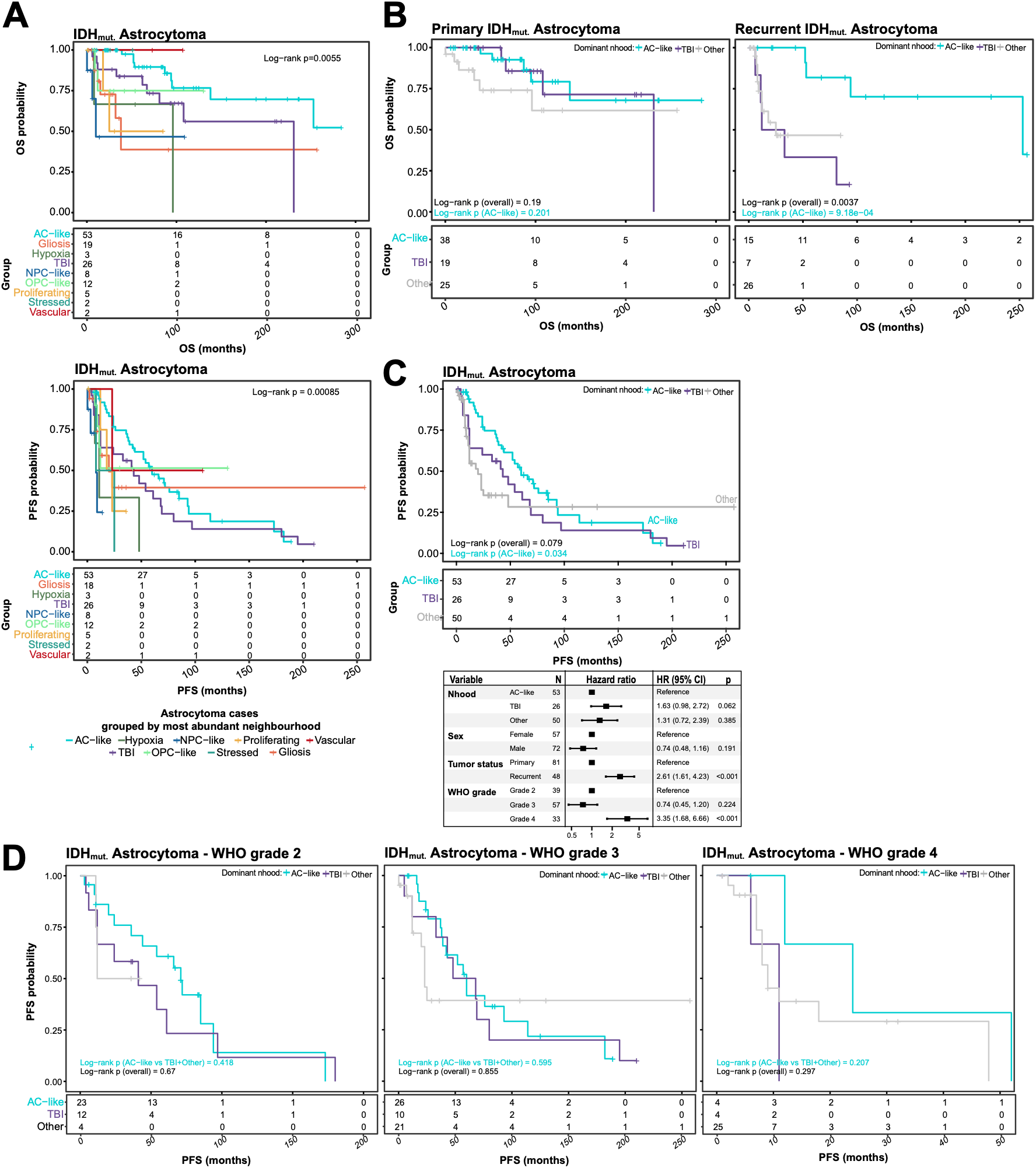
Survival of patients with IDH-mutant astrocytoma stratified by dominant neighbourhood. **(A)** Kaplan–Meier curves of overall survival (OS, top) and progression free survival (PFS, bottom) for astrocytoma patients, stratified by dominant neighbourhood. **(B)** OS in primary (left) and recurrent (right) astrocytoma, stratified by AC-like, TBI or all other dominant neighbourhoods (Other). **(C)** Kaplan–Meier curves of PFS for astrocytomas grouped by their most abundant neighbourhood: AC-like, TBI, or other neighbourhoods (Other) (n = 129 tumours from 124 patients, including 5 with both primary and recurrent tumours). Numbers at risk are shown below the curves. Forest plot: hazard ratios (HRs) with 95% confidence intervals and P-values from the multivariable Cox model. **(D)** Kaplan–Meier curves of PFS for astrocytomas grouped by their most abundant neighbourhood: AC-like, TBI, or other neighbourhoods (Other). Numbers at risk are shown below the curves. P-value: two-sided log-rank test across all three groups (overall) and for AC-like versus the other two groups combined (AC-like vs TBI+Other).

**Extended Data Figure 12.**
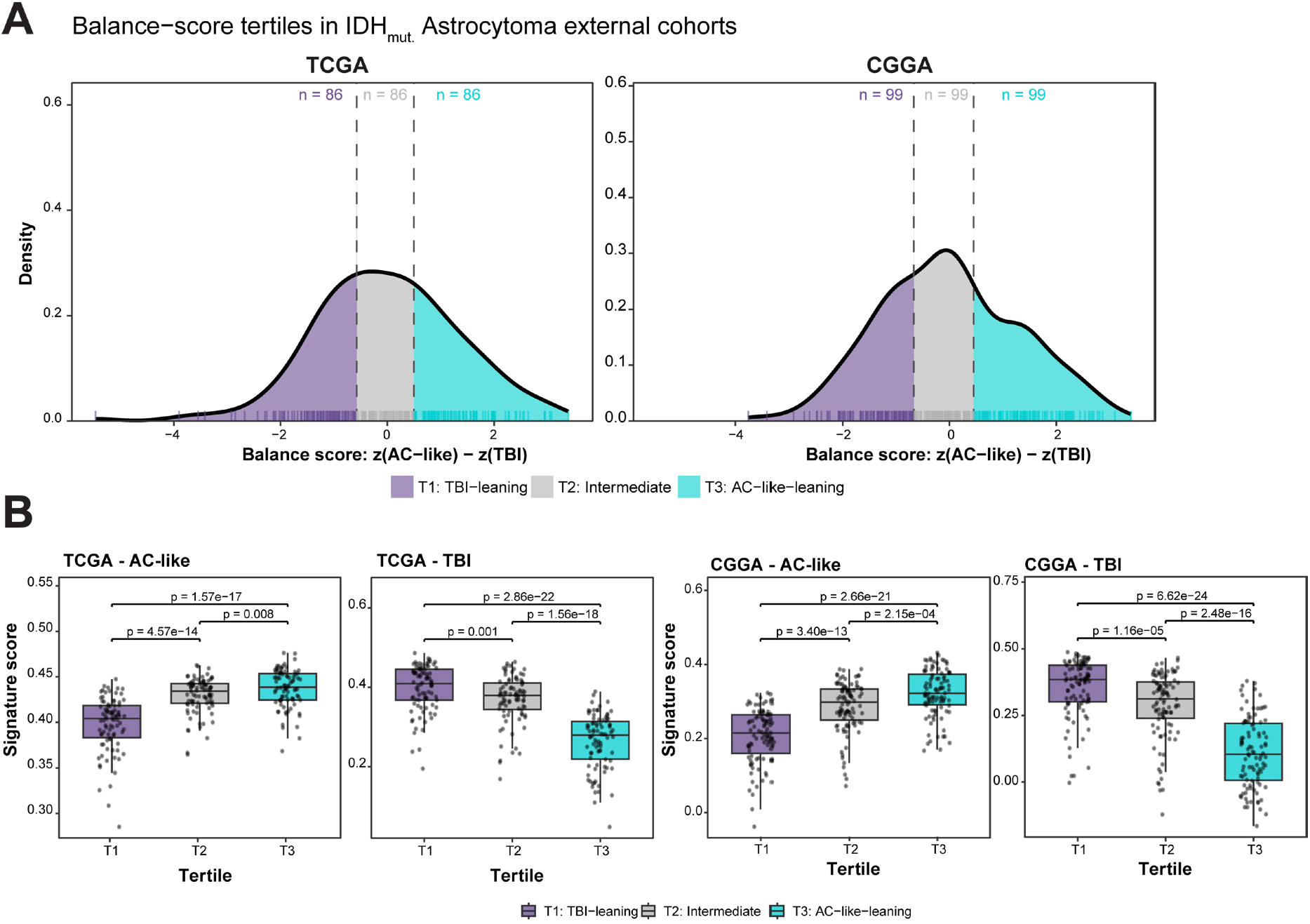
AC-like and TBI signature balance in external IDH-mutant astrocytoma cohorts. **(A)** Distribution of the balance score, defined as z(AC-like) − z(TBI), in IDH-mutant astrocytomas from TCGA (n = 258 patients) and CGGA (n = 297 patients). Curves show kernel density estimates; the rug marks individual tumours. Dashed lines indicate the tertile boundaries, computed within each cohort, which define T1 (TBI-leaning), T2 (intermediate) and T3 (AC-like-leaning) (n per tertile shown above each panel). (**B)** AC-like and TBI signature scores per balance-score tertile in TCGA and CGGA. Boxes show the median and interquartile range; whiskers extend to 1.5× the interquartile range; dots represent individual tumours. P-values: two-sided pairwise Wilcoxon rank-sum tests, Benjamini–Hochberg-adjusted within each panel. Abbreviations: AC-like: Astrocyte-like; TBI: tumour-brain resident interface.

## Acknowledgements

M.G. is supported by the Robert Bosch Foundation. A.K.S. received funding by the Emmy-Noether Programme by the Deutsche Forschungsgemeinschaft, DFG (project ID SU 1548/1-1), Hertie Network of Excellence in Clinical Neuroscience, Stiftung Sibylle Assmus, Rolf M. Schwiete Stiftung, Else Kröner-Fresenius-Stiftung (EKFS), and Stefan Kaiser Consortium for Astrocytoma Research (SKCAR). The work was supported by the SFB grant UNITE Glioblastoma (SFB1389). F.E.H. is a fellow of the Heidelberg College of Neuro-Oncology, funded by the Else Kroener Fresenius Foundation (2023_EKFK.02). F.M.I. is supported by an NCT-fellowship and is a fellow of the DKTK School of Oncology. D.T.O. is supported by European Research Council (788937), the Helmholtz Society, the Health + Life Science Alliance Heidelberg-Mannheim, and the Wilhelm Sander Stiftung. P.D.M. was supported by German Cancer Aid through the funding instrument ‘Mildred-Scheel-Doctoral Program’ (No: 70117260)

Part of the analysis was performed on data generated by the TCGA Research Network: https://www.cancer.gov/tcga.

Parts of Figures 1, 4, and 5 were created using BioRender.com under an academic publication license.

## Declaration of generative AI and AI-assisted technologies during manuscript preparation

During the preparation of this work the authors used ChatGPT (OpenAI) and Claude (Anthropic) in order to assist with data analysis and grammar and style checking. After using this tool/service, the authors reviewed and edited the content as needed and took full responsibility for the content of the published article.

## Conflict of interest

F.S. and A.v.D. is a shareholder and cofounder of Heidelberg Epignostix GmbH. F.S. also serves as an advisor to Epignostix. M.G. is a co-founder of Chronosight.ai.

## Authorship

M.G., A.K.S. and F.S. conceptualised and supervised the study. A.M. wrote the manuscript with input from S.R., M.G., A.K.S., and F.S. A.M. curated data, performed the principal analyses (spatial transcriptomics, multiplexed immunofluorescence, survival analysis, genetic associations, downstream histopathology analysis), designed the 5,096-gene spatial transcriptomics and multiplexed immunofluorescence panels, co-ordinated data generation, interpreted results, and prepared the figures. S.R. performed histopathological and survival analyses, contributed to the interpretation of results and manuscript preparation. Z.S. designed the 344-gene Xenium panel, contributed to data generation, and performed spatial transcriptomics analysis. G.R. performed cell segmentation of the Xenium dataset, developed the interactive data browser with input from A.M., and conducted multimodal alignment. M.R. provided single-nucleus RNA-seq data, performed inferCNV analysis and spatial transcriptomics analysis under the supervision of F.S. E.P. co-designed the multiplexed immunofluorescence panel and performed the corresponding experiments under the supervision of S.D. D.C. contributed to data generation and provided feedback on spatial transcriptomics analyses together with H.L. and Y.T. N.R.K. prepared tissue samples for Xenium experiments. N.W. provided technical support. M.B. supported multimodal alignment.

A.P. provided DNA methylation data. P.S. and J.P.M. supported spatial transcriptomics data generation. F.E.H., P.D.M., F.M.I., S.S., N.K., M.Ra., S.K., N.E., M.P., C.H.-M., L.B., W.W., A.W., A.v.D., and F.S. collected and provided patient samples and clinical information. A.K.S. coordinated tissue sample collection, assembled the cohort, and supervised histopathological annotation. All authors reviewed and provided feedback on the manuscript.

## Data and code availability

All processed and annotated spatial transcriptomics data objects have been deposited to Zenodo and will be made publicly available upon publication. Spatial multimodal dataset is also available at https://gliomap.dkfz.de/.

To adhere to local regulations, DNA methylation, H&E and clinical outcome data can be shared after completion of a data transfer agreement for non-commercial use. Requests should be made to the corresponding author(s), with responses expected within 14 days of receipt of request.

All code used for data analysis and figure generation is available in github (https://github.com/gerstung-lab/Panglioma_Spatial_tx). Code for multimodal registration is available in https://github.com/scOpenLab/MultimodalRegistration.

## Methods

### Ethics and inclusion statement

This study complied with ethics regulations as approved by the Ethics Committee Heidelberg (S-318/2022). Samples were collected from adult patients after obtaining written informed consent in accordance with the Declaration of Helsinki and patients were not compensated for participation. Patient sex was self-reported. No exclusions were made based on race, ethnicity, sex, age, gender or other social factors.

### NGS panel sequencing

Genomic DNA was extracted exclusively from formalin-fixed paraffin-embedded (FFPE) tissue blocks. Targeted DNA sequencing was performed at the Department of Neuropathology, University Hospital Heidelberg, using a custom-designed hybrid capture next-generation sequencing (NGS) panel. This panel encompasses the complete coding regions (all exons with flanking ±25 bp) as well as selected intronic and promoter regions of 130 genes, enabling detection of single nucleotide variants (SNVs), small insertions and deletions (indels), exonic rearrangements, and recurrent gene fusions.

### DNA methylation profiling

Genome-wide DNA methylation profiling was carried out using the Illumina Infinium MethylationEPIC BeadChip (v.1.0). Raw IDAT files were processed with the *minfi* Bioconductor package, including background and dye-bias correction prior to beta value calculation. Probes were filtered to exclude those located on sex chromosomes, containing single-nucleotide polymorphisms near or at the interrogated CpG site, non-uniquely mapping probes, and those not represented on the EPIC array v.2 design, ensuring compatibility with cross-cohort datasets. Classification of methylation profiles was performed using the Heidelberg brain tumour classifier (MNP) v.12.8 (https://www.molecularneuropathology.org/mnp/).

Uniform manifold approximation and projection (UMAP) based on the DNA methylation profiles was created using only the 32,000 probes with the highest variance across the cohort consisting of 304 profiles. The selected probes were each z-score normalised using sklearn’s StandardScaler and PCA dimensionality reduction was performed. The 94 leading principal components were used as input for the UMAP with parameters min_idst=0.4. The number of high variance probes and the number of principal components as well as the decision to standardise the probes was adopted from Capper et al. ^2^.

### Single nucleus RNA sequencing data production, preprocessing and annotation

Single-nucleus RNA sequencing (snRNA-seq) was performed on formalin-fixed paraffin-embedded (FFPE) tissue from 14 glioblastoma patients. Nuclei were isolated using the 10x Genomics Nuclei Isolation Kit (PN-1000494) following the manufacturer’s protocol. Libraries were prepared using the Chromium Single Cell Fixed RNA Kit (16-plex, PN-1000476) in combination with the Chromium Chip Q (PN-1000418) and the Dual Index Set Plate TS Set A (PN-10000251).

Sequencing was performed on an Illumina NovaSeq 6000 platform using NovaSeq 6000 S2 Reagent Kit v1.5 (100 cycles, PN-20028316) and NovaSeq 6000 S4 Reagent Kit v1.5 (200 cycles, PN-20028313) with PhiX (PN-15041963) according to the manufacturer’s recommendations.

Data preprocessing was performed using CellRanger (version 7.0.0). Downstream analysis, quality control and cell-type annotation were using Seurat (v4.3) in R (v4.3.1).

Copy number variants (CNVs) were inferred using the inferCNV package (version 1.3.3; https://github.com/broadinstitute/infercnv). A mix of all healthy cells was used as reference. The adjusted expressions of the genes in the inferCNV object were used to calculate the gains and losses of the chromosomes. For the inference a subset of roughly 10,000 healthy cells was used.

### Xenium spatial transcriptomics (custom targeted panel)

Five-micrometer sections were cut from FFPE tissue microarray (TMA) blocks of CNS tumours and prepared for 10x Genomics Xenium analysis following the FFPE tissue preparation guide (CG000578 Rev C) and Xenium in situ gene expression User Guide (CG000582, 10x Genomics). Sections were placed within the imaging area of Xenium slides, dried on a thermocycler with a dedicated adapter at 42°C for 3 h, and then dried further in a desiccator for one day. In situ expression profiling with a custom [344]-gene panel (Supplementary Table 2) was performed on the Xenium Analyzer (instrument software v3.1.0.0, analysis software xenium-3.1.0.4) by the scOpenLab.

### Spatial transcriptomics preprocessing and TME annotation

Cells were assigned to individual TMA cores using core masks generated with Segment Anything ^44^. Downstream analysis was restricted to nuclear transcripts, which were extracted separately from the output of each Xenium run. Nucleus locations were taken from the nucleus boundaries file (nucleus_boundaries.csv.gz), and transcript quality (qv column) and nuclear overlap (overlaps_nucleus column) were taken from the transcripts file (transcripts.parquet). Only transcripts with a Phred-scaled quality value (QV) above 20 that overlapped a nucleus were retained.

Cell-type labels were first assigned to a subset of high-quality cells (more than 50 transcripts per cell). Raw counts were normalised to the median total count across cells (*sc.pp.normalize_total* with default settings), log-transformed (*sc.pp.log1p*) and scaled to zero mean and unit variance per gene (*sc.pp.scale*) using scanpy ^45^. Principal component analysis was performed on all genes in the panel, and the first 30 components were retained. A 15 nearest-neighbour graph was constructed from these components using Euclidean distance. The Leiden community detection algorithm was run on this graph at resolution 2.0 and where needed subclustering was performed ^46^. The same graph was used to compute the two-dimensional UMAP visualisation.

To transfer these annotations to all cells, we trained an scVI model with a Poisson likelihood on a stratified subsample of the annotated cells, which prevents abundant populations from dominating the training ^25^. The scVI model was used to initialise an scANVI model ^47^, which was trained with the annotated cell types as labels. The trained scANVI model was then applied to the raw counts of all nuclei, and each cell was assigned the cell type with the highest predicted probability. Cells with fewer than 10 transcripts were excluded from downstream analysis.

### Sampling consistency analysis

To assess how representative a single core is of its tumour, we compared the transcriptomic profiles of cores from the same patient. Cores with fewer than 200 cells were excluded. For each core, raw nuclear counts were summed across all cells into a pseudobulk profile and converted to log-transformed counts per million (log1p(CPM)). Each gene was then standardised to zero mean and unit variance across cores. Similarity between two cores was measured as the Pearson correlation of their standardised profiles.

Core pairs were grouped into three categories: (I) cores from the same patient; (II) cores from the same patient and different primary/recurrent status (III) cores from different patients with the same diagnosis and primary/recurrent status; (IV) cores from different patients that differed in diagnosis or status.

Pairs that share a core are not independent, so differences between groups were assessed with permutation tests (2,000 permutations) on the difference in median correlation. For the comparison of same-patient pairs with different-patient pairs of matching diagnosis and status, patient labels were permuted among cores within each diagnosis–status stratum. For the comparison of matching with non-matching different-patient pairs, diagnosis–status labels were permuted between patients, keeping each patient’s cores together. One-sided p-values were calculated as (number of permuted differences ≥ observed difference + 1)/(number of permutations + 1). For each same-patient pair, we also report the percentile of its correlation among different-patient pairs with the same diagnosis and status.

### Tumour states annotation

To identify recurrent transcriptional programs across tumour cells, we used a linearly decoded variational autoencoder (LDVAE; scvi-tools LinearSCVI) ^24^ with a modified, NMF-style decoder. In this decoder, each program is a non-negative distribution over genes, and each cell’s expression is a non-negative combination of program activities. Methylation class was included as the batch variable and modeled as an additional program with a fixed logit offset (batch_logit = 1.0). The model used 9 latent dimensions and a Poisson likelihood. It was trained for 400 epochs on a 1% subsample of tumour cells, stratified by diagnosis, and then applied to all tumour cells. The *XIST* gene was excluded.

### Spatial graph-based identification of cellular neighbourhoods

To characterise the spatial organisation of tumour states within the tumour microenvironment, we identified cellular neighbourhoods. A spatial neighbor graph of all cells was constructed from cell centroids as vertices using Delaunay triangulation from squidpy (v1.6.1) ^48^, with each TMA placed at a separate offset in coordinate space, and edges longer than 50 µm were removed.

For each cell, we computed a neighbourhood profile from the predictions of the cell-type scANVI model, consisting of the cell-type composition of the cell and its first-degree neighbors, concatenated with that of its second-degree neighbors. Two versions of this profile were used. The probability-based version averages the predicted cell-type and tumour state probabilities, and the count-based version counts cells by their highest-probability cell type. To define neighbourhoods, we sampled up to 100 cells from each core, built a k-nearest-neighbor graph between these cells on the probability-based profiles (scanpy defaults, k = 15) and clustered it with the Leiden algorithm at resolution 0.5 ^46^. The resulting clusters were then used as labels to train an scVI model with a Poisson likelihood on the count-based profiles, followed by an scANVI model ^47^. The trained scANVI model then predicted the neighbourhood of every cell in the full dataset from its count-based profile.

### Spatial neighbourhood interaction analysis

Spatially contiguous cells assigned to the same neighbourhood were grouped into connected regions within each tissue core, similarly to our previous work ^49^. Regions containing fewer than 15 cells were excluded. Spatial contacts between regions of different neighbourhoods were quantified using the spatial connectivity Delaunay graph, with each unique region–region contact counted once.

Expected neighbourhood–neighbourhood contact frequencies were estimated independently for each core using 10,000 spatial permutations that preserved the core-specific spatial organisation. Enrichment or depletion of observed contacts relative to permutation-derived expectations was assessed using Poisson generalised linear models with the log expected contact count included as an offset. Exponentiated model coefficients were reported as rate ratios, with values >1 indicating enrichment and <1 indicating depletion.

Diagnosis-specific interaction profiles were obtained by fitting the same model separately to the cores of each diagnostic group, using the core-level expected counts described above. To compare neighbourhood-pair repertoires between diagnoses, a neighbourhood pair was considered present in a diagnosis if it was assessable (non-zero expected contact count) in at least two cores of that diagnosis and yielded a finite rate-ratio estimate. Similarity between two diagnoses was quantified as the Jaccard index, defined as the number of neighbourhood pairs present in both diagnoses divided by the number of pairs present in either.

### Association of genetic and molecular alterations with neighbourhood abundance

For each patient, the fraction of cells assigned to each spatial neighbourhood was calculated. Associations between neighbourhood fractions and genetic or molecular alterations were tested separately for each neighbourhood, comparing tumours with and without each alteration using two-sided Welch’s t-tests. Before testing, neighbourhood fractions were logit-transformed. To avoid undefined values, a pseudocount ε was applied: fractions of exactly 0 or 1 were replaced by ε or 1 − ε, respectively, where ε was half of the smallest non-zero fraction of that neighbourhood among the tumours included in the comparison. P-values were corrected for multiple testing across neighbourhoods within each comparison using the Benjamini–Hochberg procedure, and q < 0.1 was considered significant. Comparisons were performed only when both groups contained at least 3 tumours.

### Histology feature extraction and clustering

Consecutive H&E-stained sections corresponding to the Xenium samples were scanned using a Leica Aperio AT2 scanner at 0.25mpp (40x magnification). The images were segmented and tiled using the TRIDENT library (v. 0.2.0; https://github.com/mahmoodlab/TRIDENT) ^50^. The feature files that were produced for each slide were separated into files for each individual TMA core. The TITAN ^31^slide-level pathology foundation model was used to generate a 768-dimensional vector representation of each TMA core’s H&E image.

The two-dimensional UMAP visualisation of these features shown in Figure 4 was generated using the settings ‘metric=”cosine”’ (umap-learn 0.5.12) from the 768-dimensional TITAN features without prior standardisation or dimensionality reduction.

The histology clusters were derived from the TITAN features by creating a 15-nearest neighbour graph based on cosine similarity on which the Leiden community detection ^46^ algorithm was run at resolution 0.5 through the scanpy package (1.12.1) ^45^.

### Association of histology clusters with diagnosis and neighbourhood composition

Analyses were restricted to astrocytoma low-grade, astrocytoma high-grade and glioblastoma, in which histology clusters spanned multiple diagnoses (206 cores, 177 patients). neighbourhoods present at >1% in fewer than 5% of cores were excluded. Histology cluster membership was predicted by multinomial logistic regression (scikit-learn v. 1.7.2; L2 penalty, C = 1) using either diagnosis (one-hot encoded) or neighbourhood fractions as predictors, z-scored within each training fold. Performance was estimated by 5-fold cross-validation stratified by cluster, with all cores of a patient in the same fold, repeated 20 times. One-vs-rest AUCs were computed per cluster from pooled out-of-fold predictions and averaged across clusters (macro-AUC).

To test whether neighbourhoods are associated with clusters independently of diagnosis, the improvement in cross-validated log loss from adding neighbourhoods to diagnosis was compared with a null distribution obtained by permuting neighbourhood profiles among cores of the same diagnosis (200 permutations); p = (1 + number of permutations ≥ observed) / (1 + 200). To compare the contributions of diagnosis and neighbourhoods, a joint multinomial logistic regression including both (all predictors z-scored) was fitted on all cores. Exponentiated coefficients give the odds ratio of each cluster, relative to the average across clusters, per SD increase of each predictor. 95% confidence intervals and two-sided p-values were obtained from 500 patient-level bootstrap resamples and corrected across all cluster–predictor pairs using the Benjamini–Hochberg procedure.

### Same-slide spatial transcriptomics and multiplexed immunofluorescence

Same-slide spatial transcriptomics and multiplexed immunofluorescence were performed as previously described ^51^. Five-micrometer sections of 16 low-grade astrocytoma tissue cores were profiled using the Xenium Prime 5K gene panel with a 96-gene custom add-on, also used for profiling of lymphoma ^52^ (**Supplementary Table 5**; 10x Genomics; instrument software v3.1.0.0, analysis software xenium-3.1.0.4). Sections were prepared as described for the custom-panel Xenium analysis above.

After Xenium data acquisition, the same slides were used for multiplexed immunofluorescence, according to Sivajothi et al. 2024. Tissue was kept in 50% glycerol in PBS storage solution at 4 °C for up to 2 weeks. Heat-induced antigen retrieval was performed for 20 min at 155–160 °C in Tris–EDTA buffer (pH 9). Sections were incubated overnight at 4°C with a cocktail of DNA oligonucleotide-conjugated antibodies (**Supplementary Table 6**), conjugated using previously published oligonucleotide sequences ^53^. After incubation, tissue was fixed in three steps using 1.6% paraformaldehyde (PFA), 100% methanol and bis(sulfosuccinimidyl) suberate (BS3).

Imaging was performed on the PhenoCycler-Fusion system (Akoya Biosciences) according to the manufacturer’s instructions, using Atto550- and Atto647N-labeled complementary DNA barcodes for cyclic imaging. Raw files were processed into a single qpTIFF file with the Fusion software (v2.4.0). For each cycle, the nuclear stain was compared with the reference cycle (by default the DAPI signal of the second cycle), and the resulting offsets were used to align all cycles by rigid translation. Background was subtracted using blank cycles at the beginning and end of each run, deconvolution was carried out with the Microvolution package, and shading was corrected with BaSiC. Adjacent tiles were registered using the Microscopy Image Stitching Tool library. Further details are given in the manufacturer’s technical documentation (https://help.codex.bio/codex/processor/technical-notes).

### Multimodal image registration and spatial transcriptomics annotation

Individual TMA cores were segmented using Segment Anything (ViT-H, sam_vit_h_4b8939 checkpoint)^44^, and alignment was performed at the core level. Xenium and multiplexed immunofluorescence images were registered at the pixel level on the basis of the DAPI signal. Scale-invariant feature transform (SIFT) keypoints were detected and matched with OpenCV (v4.11.0; FLANN matcher, Lowe’s ratio test), and the matched landmarks were used to estimate an affine transformation by least squares (scikit-image v0.25.2, via SpatialData v0.3.0). The aligned data were stored in the SpatialData format ^54^ and analyzed as described for the custom-panel dataset.

Cell types and tumour states in the high-plex spatial transcriptomics cohort were annotated as described above. Spatial neighbourhood annotations were transferred from the discovery cohort using multinomial logistic regression implemented in scikit-learn (v1.7.2). Cell-type annotations were harmonised between datasets, and local neighbourhood composition was quantified using cell-type counts derived from Delaunay connectivity graphs. These profiles were used to train the classifier on annotated low-grade astrocytomas from the discovery cohort and predict neighbourhood identities in the independent cohort.

### Neighbourhood gene signatures and scoring in bulk transcriptomics

To create gene expression signatures for each neighbourhood which could be used as surrogates in bulk expression measurements, pseudobulk profiles were generated from spatial transcriptomics cells. For each neighbourhood and each core the expression counts were added together (decoupler v. 2.1.6; ‘<u>decoupler.pp</u>.pseudobulk with sample_col=’core’, groups_col=’nhood’) ^55^. Each core might thus give rise to as many pseudobulks as there are neighbourhoods. Pseudobulk profiles that consist of the expression of fewer than 30 cells were excluded. The remaining profiles were normalised to each sum to 100 (<u>scanpy.pp</u>.normalize_total with target_sum=100).

For each neighbourhood a Wilcoxon’s rank-sum test was performed to identify genes that differ in expression between pseudobulks from that neighbourhood as compared to all other neighbourhoods (<u>sc.tl</u>.rank_gene_groups method=’wilcoxon’, reference=’rest’). The 10 genes with the lowest p-values that show an increase in expression were included in the signatures. The table of signature genes is Supplementary Table 8. Signatures were derived from the entire discovery cohort. Scoring the signatures in bulk transcriptomics samples was done using singscore (R 4.3.3, singscore v. 1.22.0) ^56^.

### Validation survival analysis

For validation of spatial transcriptomics-derived features, we assembled two previously published, clinically annotated bulk transcriptomics datasets of IDH-mutant astrocytomas from The Cancer Genome Atlas (TCGA) ^57,58^ and the Chinese Glioma Genome Atlas (CGGA) ^34^. TCGA: The TCGA-GBM and TCGA-LGG cohorts were combined and restricted to IDH-mutant astrocytomas using the revised sample annotation in accordance with the WHO 2021 CNS tumour classification guidelines provided by Leiria de Mendonça et al. ^33^. https://github.com/sysbiomed/MONET/blob/master/Final-outputs/SIMPLIFIED_CLASSIFICATION_TCGA_2016_2021.csv; commit 4361ed6.

Gene expression data (log2(TPM + 1); v. 05-10-2024) were downloaded from the UCSC Xena Browser (https://xenabrowser.net/datapages/) on 9 February 2026. Signatures were scored as described above. Where several samples were available per patient, one sample was used, with the primary tumour preferred. Curated overall survival (OS) data were taken from the TCGA Pan-Cancer Clinical Data Resource ^59^.

CGGA: The mRNAseq_693 and mRNAseq_325 datasets (CGGA.mRNAseq_693.RSEM-genes.20200506.txt and CGGA.mRNAseq_325.RSEM-genes.20200506.txt) and the corresponding clinical data were downloaded from https://www.cgga.org.cn/download.jsp on 24 June 2026. Expression values were converted to TPM and limited to genes shared by both datasets. The expression counts for genes with the same gene symbol were added together. IDH-mutant astrocytomas were selected based on the provided clinical annotation (IDH mutation status “Mutant” and 1p/19q status “Non-codel”).

Gene signatures for the AC-like and TBI neighbourhoods were derived from the discovery spatial transcriptomics dataset as described above. Scores were z-scored within each cohort, and a balance score was calculated as z(AC-like) − z(TBI), with positive values indicating relatively higher AC-like than TBI expression. tumours were divided into tertiles of the balance score within each cohort (T1, TBI-leaning; T2, intermediate; T3, AC-like-leaning).

Survival across these tertiles was compared using Kaplan–Meier estimates and a two-sided log-rank test, with pairwise log-rank tests adjusted using the Benjamini–Hochberg method. Signature scores were compared between tertiles using pairwise two-sided Wilcoxon rank-sum tests with Benjamini–Hochberg adjustment. Analyses were performed in R (v. 4.5.1) using the survival (v. 3.8.3) and survminer (v. 0.5.0) packages. Survival times were converted from days to months (days/30.4375), and the full follow-up was used.

### Statistics and reproducibility

No statistical method was used to predetermine the sample size. No data were excluded from the analyses. Data collection and analysis were not blinded to the conditions of the experiments. Statistical tests used for individual analyses are described in the figure legends and the Methods.

## Notes

### Competing Interest Statement

The authors have declared no competing interest.

### Author Declarations

This study complied with ethics regulations as approved by the Ethics Committee Heidelberg (S-318/2022). Samples were collected from adult patients after obtaining written informed consent in accordance with the Declaration of Helsinki and patients were not compensated for participation.

